# Modeling serological testing to inform relaxation of social distancing for COVID-19 control

**DOI:** 10.1101/2020.04.24.20078576

**Authors:** Alicia N.M. Kraay, Kristin N. Nelson, Conan Y. Zhao, David Demory, Joshua S. Weitz, Benjamin A. Lopman

## Abstract

Serological testing remains a passive component of the current public health response to the COVID-19 pandemic. Using a transmission model, we examined how serology can be implemented to allow seropositive individuals to increase levels of social interaction while offsetting transmission risks. We simulated the use of widespread serological testing in three metropolitan areas with different initial outbreak timing and severity characteristics: New York City, South Florida, and Washington Puget Sound. In our model, we use realistic serological assay characteristics, in which tested seropositive individuals partially restore their social contacts and act as immunological ‘shields’. Compared to a scenario with no intervention, beginning a mass serological testing program on November 1, 2020 was predicted to avert 15,000 deaths (28% reduction, 95% CrI: 0.4%-30.2%) in New York City, 3,000 (31.1% reduction, 95% CrI: 26.4%-33.3%) in South Florida and 10,000 (60.3% reduction, 95% CrI: 50.2%-60.7%) in Washington State by June 2021. In all three sites, widespread serological testing substantially blunted new waves of transmission. Serological testing has the potential to mitigate the impacts of the COVID-19 pandemic while also allowing a substantial number of individuals to safely return to social interactions and economic activity.

**Significance:** The SARS-CoV-2 pandemic led many countries to implement broadly impactful social distancing interventions. In the U.S., many measures are still partially in place. While these policies slowed the epidemic, they also caused substantial social and economic disruption. Here, we explore how using available serological assays to inform the individualized relaxation of social distancing could allow a gradual return to more normal levels of economic and social activity while mitigating public health risk. We find that large scale serological testing in major metropolitan areas has the potential to lower expected COVID-19 deaths, reduce peak health system burden to manageable levels, and also allow social distancing interventions to be substantially relaxed.

## Introduction

SARS-CoV-2 emerged in China in late 2019 leading to the pandemic of COVID-19, with over 50 million detected cases and over 1.2 million deaths globally as of November 9, with nearly 10 million detected cases and 237,000 deaths reported in the U.S. by that same date (1). Unprecedented social distancing measures were enacted in early 2020 to reduce transmission and thereby blunt the epidemic peak (i.e. “flatten the curve”). In early March, U.S. states began to close schools, suspend public gatherings, and encourage employees to work from home if possible. By mid-April, 95% of the U.S. (2) and over 30% of the global population were under some form of shelter-in-place order (3). Federal social distancing guidelines expired on April 30, 2020; in late April and May, many state and local governments relaxed stay-at-home orders partially or completely to move towards ‘re-opening’ (4).

Relaxing these initially effective social distancing policies resulted in increased contacts and community transmission, and case counts began to increase as states further relaxed restrictions on public gatherings, restaurant dining, and operation of businesses (5). However, many workplaces remain completely or partially closed, schools have transitioned to some level of virtual instruction, and many businesses remain only partly operational. With the goal of maintaining the reproductive number at or less than one, public health efforts advocate a gradual return to such activities together with enhanced hygiene and mask-wearing, while maintaining stricter distancing measures for individuals at higher risk (6).

In addition to these non-specific measures, a ‘shielding’ strategy could aim to identify and deploy recovered (and likely immune) individuals as focal points for sustaining less risky interactions via ‘interaction substitution’. This strategy has the objective of sustaining interactions necessary for the functioning of essential goods and services while reducing the risk of exposing individuals who remain susceptible to infection. As the basis for a ‘shielding’ strategy, widespread serological testing programs have the potential to identify individuals or groups who are likely immune, allowing some individuals to return to activities while keeping deaths and hospital admissions at sufficiently low levels. In this strategy, individuals who test positive would preferentially replace susceptible individuals in close-contact interactions, such that more contacts are between susceptible and immune individuals rather than between susceptible and potentially infectious individuals (7).

Recent serosurveys of SARS-CoV-2 in the U.S. vary in their estimates of seroprevalence but collectively suggest that infections far outnumber documented cases (8–12). To the extent that antibodies serve as a correlate of immunity, serological testing may be used to identify protected individuals (13). While our understanding of the immunological response to SARS-CoV-2 infection remains incomplete, the vast majority of infected individuals seroconvert (14), with detectable antibody levels persisting at least several months after infection (15). Few reinfections with SARS-CoV-2 have been documented, suggesting that, while possible, reinfection is rare. Together, these data suggest that recovered individuals have substantial protection against subsequent reinfection. Once identified, antibody test-positive individuals could return to pre-pandemic levels of social interactions and therefore dilute (via ‘serological shielding’) potentially risky interactions between susceptible and infectious individuals (7).

Such strategies, however, rely on correctly identifying immune individuals. There are currently more than 50 serological assays for detection of SARS-CoV-2 antibodies that have been approved for emergency use by the Food and Drug Administration (16) with others currently in development and approved in other countries (17). The performance of these tests varies considerably (16,18,19). For the purpose of informing safe social distancing policies, specificity rather than sensitivity is of primary concern. An imperfectly specific test will result in false positives, leading to individuals being incorrectly classified as immune. If used as a basis to relax social distancing measures, there is concern that this error could heighten risk for individuals who test positive and lead to an increase in community transmission. For this reason, this paper evaluates the integration of serological testing into a COVID-19 transmission model to evaluate the level of serological testing needed to reduce expected fatalities while increasing the fraction of focal populations from social distancing.

### Model overview

To evaluate the epidemiological consequences of using mass serological testing to inform the relaxation of social distancing measures, we modeled transmission dynamics and serological testing for SARS-CoV-2 using a deterministic, compartmental SEIR-like model. Recovered, susceptible, latently infected, and asymptomatic test positive at rates that are functions of testing frequency, sensitivity (for recovered individuals), and specificity (for non-immune individuals). (Figure 1A) We model contacts at home, work, school, and other locations among three age groups: children and young adults (<20 years), working adults (20-64 years), and elderly (65+ years). We used a Markov Chain Monte Carlo (MCMC) approach to fit the model to time series of deaths (20) and cross-sectional seroprevalence (12) data from three U.S. metropolitan areas with distinct COVID-19 epidemic trajectories: the New York City Metro Region, South Florida, and the Washington Puget Sound region.

**Figure 1.**
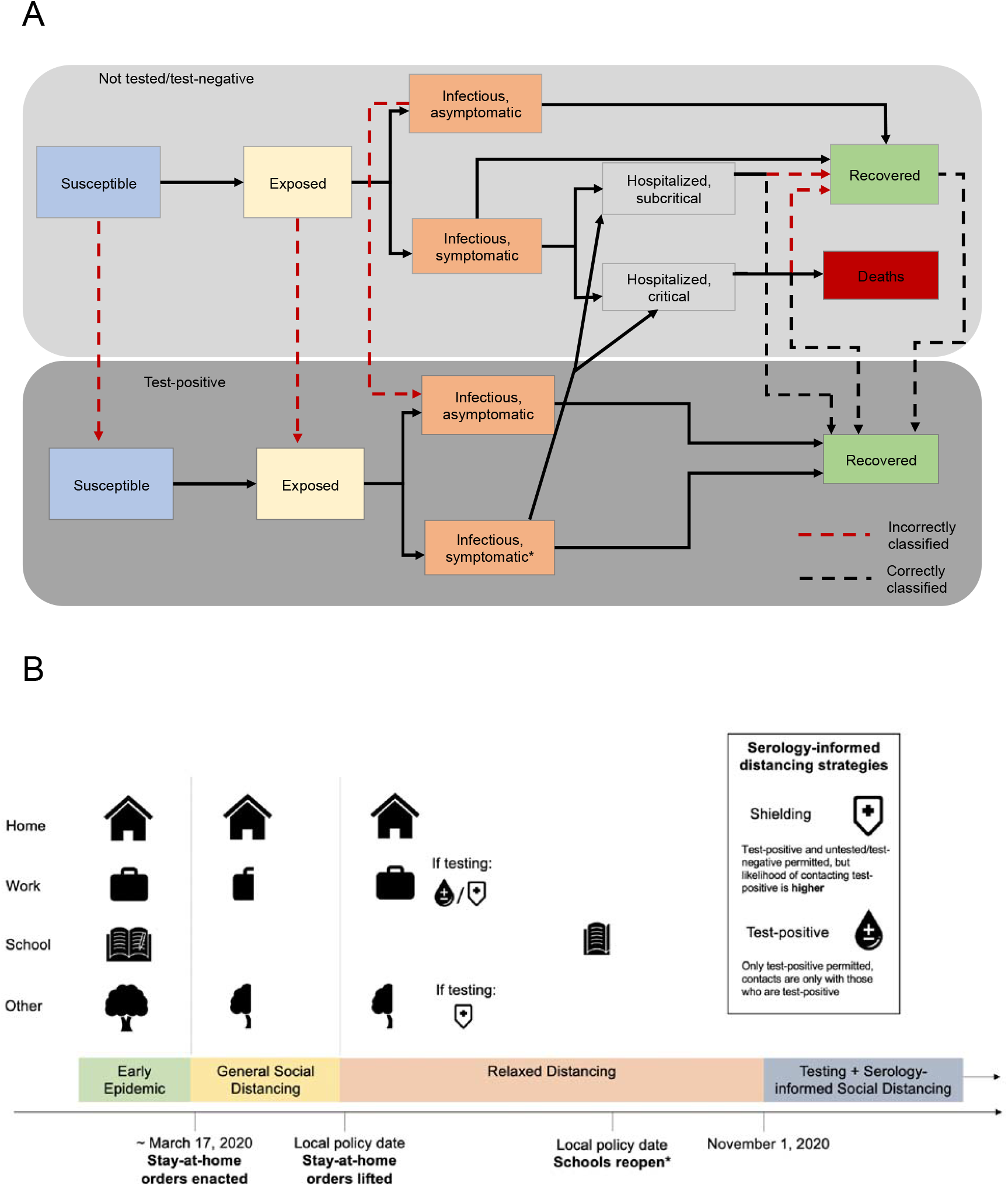
Methods diagram overview A) Overall model diagram. Serological antibody testing is shown by dashed arrows. Red dashed arrows indicate either false positives (i.e., someone is not immune, but is moved to the test-positive group) and occur at a rate that is a function of 1-specificity, or false negatives (i.e., someone is recovered, but stays in the test-negative group). True positives occur at a rate that is a function of the sensitivity. *Symptomatic infections in the test-positive group have similar severity to symptomatic infections in the not tested/test-negative group, but symptoms are not recognized as being caused by SARS-CoV-2 unless they are severe enough to warrant hospitalization. Note that the hospitalization compartments are located in the ‘Not tested/test-negative’ layer for simplicity, though individuals who incorrectly test positive could move to these compartments after developing symptomatic infection. B) All three municipalities enacted social distancing regulations in mid-March (21–23). Under these measures, we assume all contacts at school and daycare were eliminated and contacts outside of home, work, and school (‘other’) locations were substantially reduced (24) while contacts at home remained unchanged, with distancing starting at the time that stay-at-home orders were enacted in each site. (Figure 1A) We used the ‘reopening’ date, or date that stay-at-home orders were lifted in each location, as the time at which social distancing was partially relaxed. In accordance with school reopening plans in each location, we assume that schools and daycares remained closed until September 1, 2020 in South Florida and October 1, 2020 in Washington and New York, after which they reopen at 50% capacity. This 50% reduction is meant to capture that interactions in schools in Fall 2020 are substantially reduced from pre-pandemic levels due to the combination of online instruction as well as physical distancing and mask-wearing for students attending in-person. We assumed a serological testing and shielding strategy starts on November 1, 2020.

## Results

### Model Fits to Fatalities and Serological Data

We explored the impacts of social distancing on epidemic outcomes in the absence of serological testing. To do so, we first fit the model to reported deaths and to seroprevalence point estimates from each of three metropolitan areas. Model fits for each location converged and closely reproduced reported death trends through June 2020 and also matched seroprevalence estimates early in the outbreak (see Figure 2 for fits; Figures S1-S10 for full model diagnostics). We note that the probability of infection per contact and the fraction of symptomatic cases had narrow credible intervals (CrI), indicating posterior confidence in the ability of the model to uniquely identify parameter sets consistent with key features of infection and transmission. While mixing parameters exhibit parameter identifiability concerns and additional data would be needed to disentangle these parameters, the robustness of fits implies that the model outcomes early in the epidemic are insensitive to variation in these parameters – enabling us to evaluate baseline predictions with and without serological testing.

**Figure 2.**
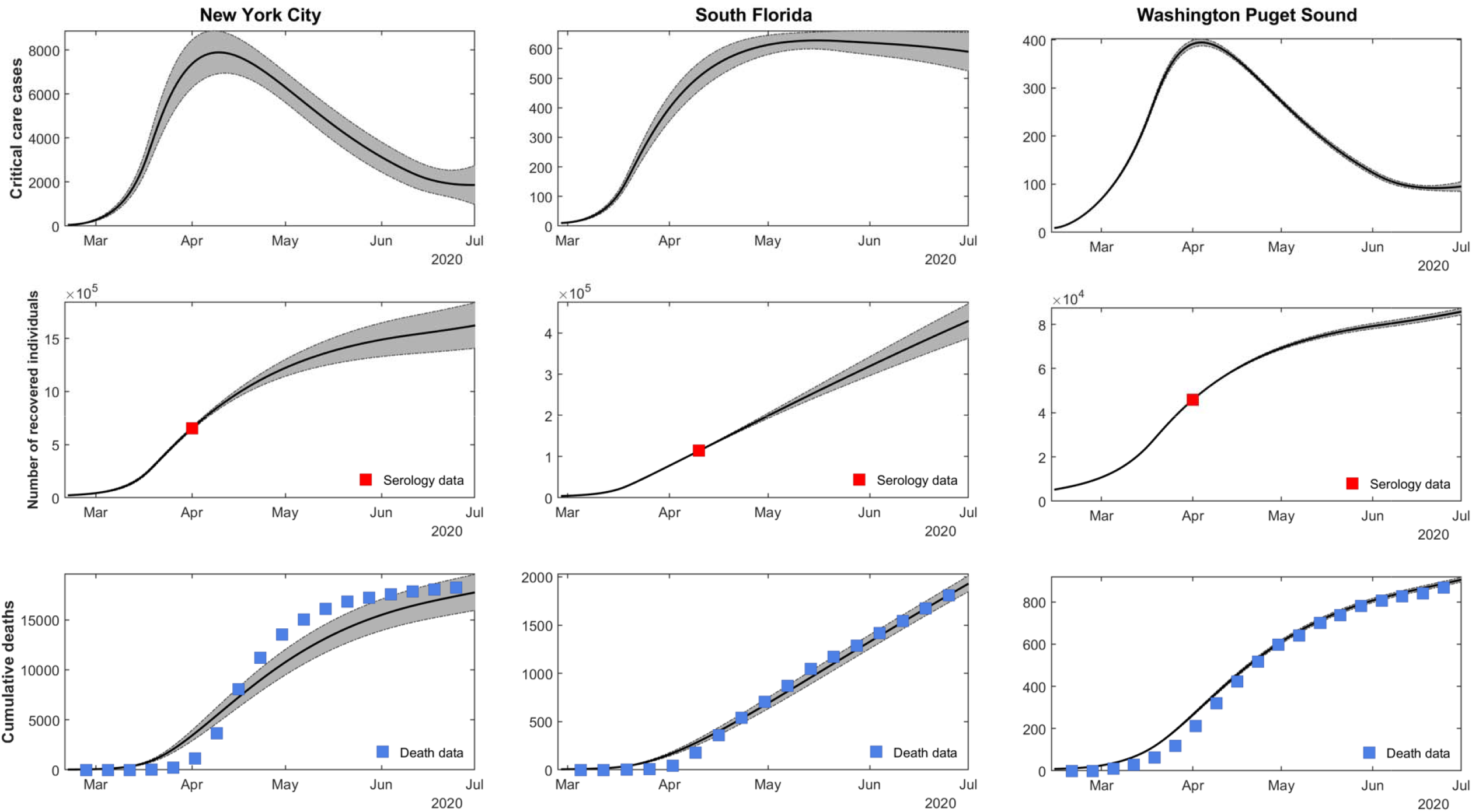
The consistency between the fitted model and the deaths/seroprevalence data for New York City, South Florida, and Washington Puget Sound. Daily critical care cases through July 1 are shown in the first row. In the second row, the cumulative number of recovered (previously infected) individuals is shown. A red square shows the seroprevalence estimates from Havers *et al*. in each location (12). In the third row, the cumulative deaths are shown, with death data shown in blue squares (20). Grey bands show 95% credible intervals, derived from the last 5,000 iterations of converged MCMC chains (ten chains from New York City and South Florida, nine chains from Washington Puget Sound).

### Epidemic dynamics in the absence of serological testing

In all three sites, our models predict a second epidemic peak in the fall and winter of 2020-2021. (Figure 3) For New York, the second peak is predicted to be smaller than the first, whereas the second wave is expected to be larger than the first wave in Washington and South Florida. If social distancing is sustained at current levels without any further interventions, our model predicts that 30-45% of the population across the three metropolitan areas (45% in New York City, 95% credible interval (CrI): 18-66%; 30% in South Florida, 95% CrI: 25-33%; and 37% in Washington, 95% CrI: 18-43%) will be infected with SARS-CoV-2 by June 2021, resulting in 79,000 cumulative deaths across the three sites (52,000 deaths in New York City, 95% CrI: 20,500-84,400; 9,200 deaths in South Florida, 95% CrI: 7,800-10,500; and 17, 000 deaths in Washington, 95% CrI: 8,100-22,500) since the start of the pandemic. (Figure 4, top row)

**Figure 3.**
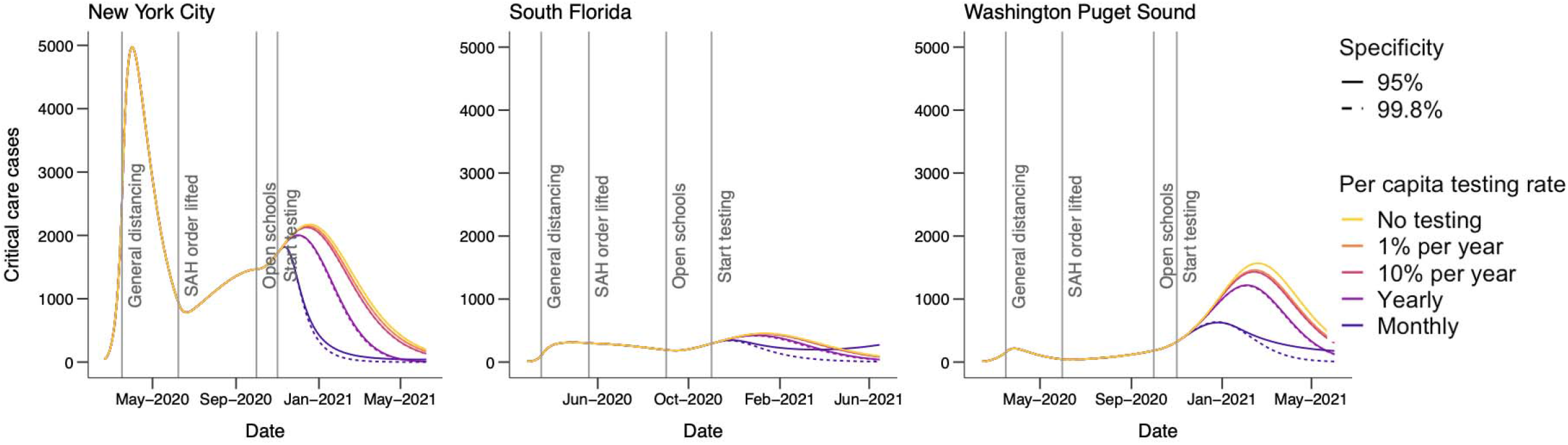
Critical care cases over time by testing level (colors) and location (panels) assuming 5:1 shielding. Dates corresponding to the start of general social distancing in March 2020 and ‘reopening’ or lifting at stay-at-home (SAH) orders in May and June 2020, are based on the dates that policies were enacted, or restrictions lifted, in each location. We assume that schools reopen at 50% capacity on September 1, 2020 in South Florida and October 1, 2020 in Washington and New York. Dotted lines show the impacts of a test with 96% specificity and solid lines show a test with 99.8% specificity.

**Figure 4.**
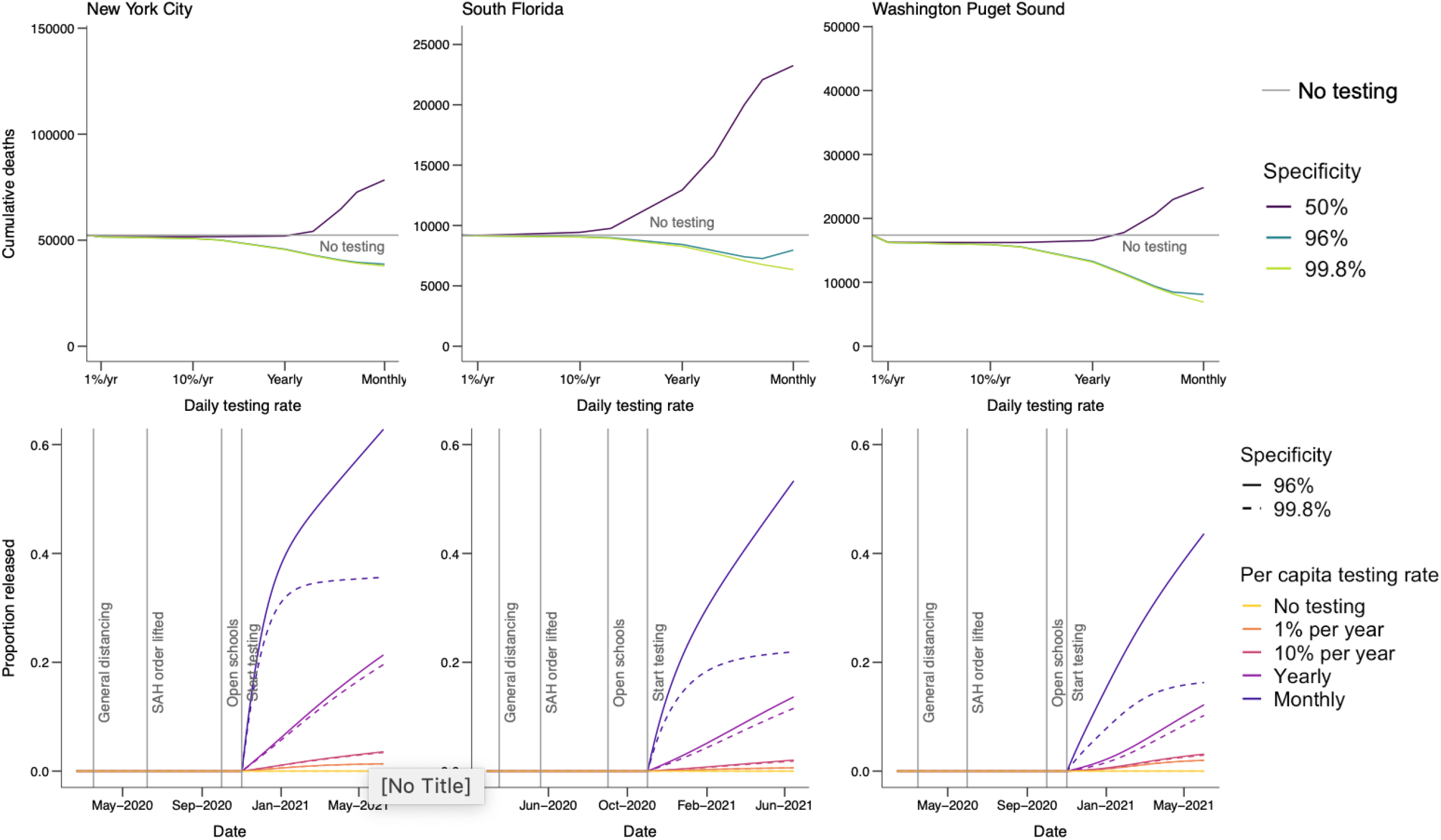
The top row shows cumulative deaths by location (panels) from March 2020 to February 2021 for the scenario with 5:1 shielding, assuming schools reopen on September 1, 2020 in South Florida and October 1, 2020 in Washington and New York. Colored lines show test specificity. if testing begins on November 1, 2020 and dotted lines show expected impact if testing had begun at ‘reopening’, or when stay at home orders were initially relaxed. The grey horizontal line shows the number of deaths in the no-testing scenario for each location. The bottom row shows the fraction of the population of each metropolitan area released from social distancing after by June 1, 2021, assuming 5:1 shielding. Line colors correspond to testing levels; blue is monthly testing (10 million tests/day) of the US population. Dashed lines show expected results with a highly specific test (specificity=99.8%) and solid lines show expected results with a test with 96% specificity.

### Epidemic dynamics with serological shielding

Next, we evaluated the benefit of serological shielding by considering the impacts of monthly testing in each metropolitan area such that test-positive individuals increase their relative rate of interactions, thereby shielding susceptible individuals and reducing the risk of transmission. Specifically, individuals who test positive return to work and increase other contacts to normal levels. We assume that test-negative/untested individuals continue to work from home if their job allows them to do so (Figure 1B). To reflect the placement of test-positive individuals in high-contact roles, we assume that contacts at work and other (non-home, non-school) locations are preferentially with test-positive persons. When shielding is 5:1 (moderate shielding) the probability of interacting with a test-positive individual is 5 times what would be expected given the frequency of test-positive individuals in the population, following the model of ‘fixed shielding’ (described in (7)). In each site, serological testing leads to a flattened epidemic curve in the fall and winter of 2020-2021. Widespread serological testing combined with moderate serological shielding (5:1) starting on November 1, 2020, using a highly specific test, could reduce cumulative deaths by June 2021 by 35% across the three sites combined. The strongest reductions are in Washington (60%, 95% CrI: 50-61%), with a lower relative impact in New York City (28%, 95% CrI: 0-30%) and South Florida (31%, 95% CrI: 26-33%). (Figure 4, top row)

### Impacts of serological testing frequency on epidemic outcomes and release from social distancing

The magnitude of the benefit from serological shielding depends on the frequency of testing, with more frequent testing resulting in both larger reductions in deaths and in a greater proportion of the population being released from social distancing if a highly specific test is used. (Figure 4, bottom row) In New York, monthly population testing is needed to maximize the potential benefit, leading to 35% of the population being released from social distancing after one year (95% CrI: 20%-68%) and deaths being reduced by 14,500 (95% CrI: 100-15,600). In contrast, annual population testing would be expected to release 19% of the population from social distancing (95% CrI: 8-35%) with 6,800 deaths averted (95% CrI: 0-6,700). More frequent testing was also beneficial in Washington; monthly testing was expected to release 16% of the population from social distancing before June 1 (95% CrI: 9-20%) with 10,000 deaths averted (95% CrI: 4,00-13,000), compared with only 10% released (95% CrI: 4-14%) and 4,000 deaths averted (95% CrI: 1,500-5,200) with annual testing. In South Florida, 22% (95% CrI: 19-25%) of the population would be released from social distancing with monthly testing, compared with 11% (95% CrI: 9-13%) with yearly testing. Monthly testing was expected to avert 2,900 deaths in South Florida (95% CrI: 2,100-3,300), whereas annual testing would avert 900 deaths (95% CrI: 700-1,100).

While increasing the intensity of social distancing towards the level of restrictions observed in April could help reduce deaths, these same benefits could be achieved by adding serological testing as part of a control strategy, allowing social distancing to be safely relaxed. As social distancing measures are relaxed further, testing frequency must increase to minimize deaths and maximize the proportion of the population that can be released (Figure 5). The extent to which testing frequency must increase to compensate for relaxing social distancing varies by location. In New York City and South Florida, distancing can be relaxed fully if monthly testing is employed.

**Figure 5.**
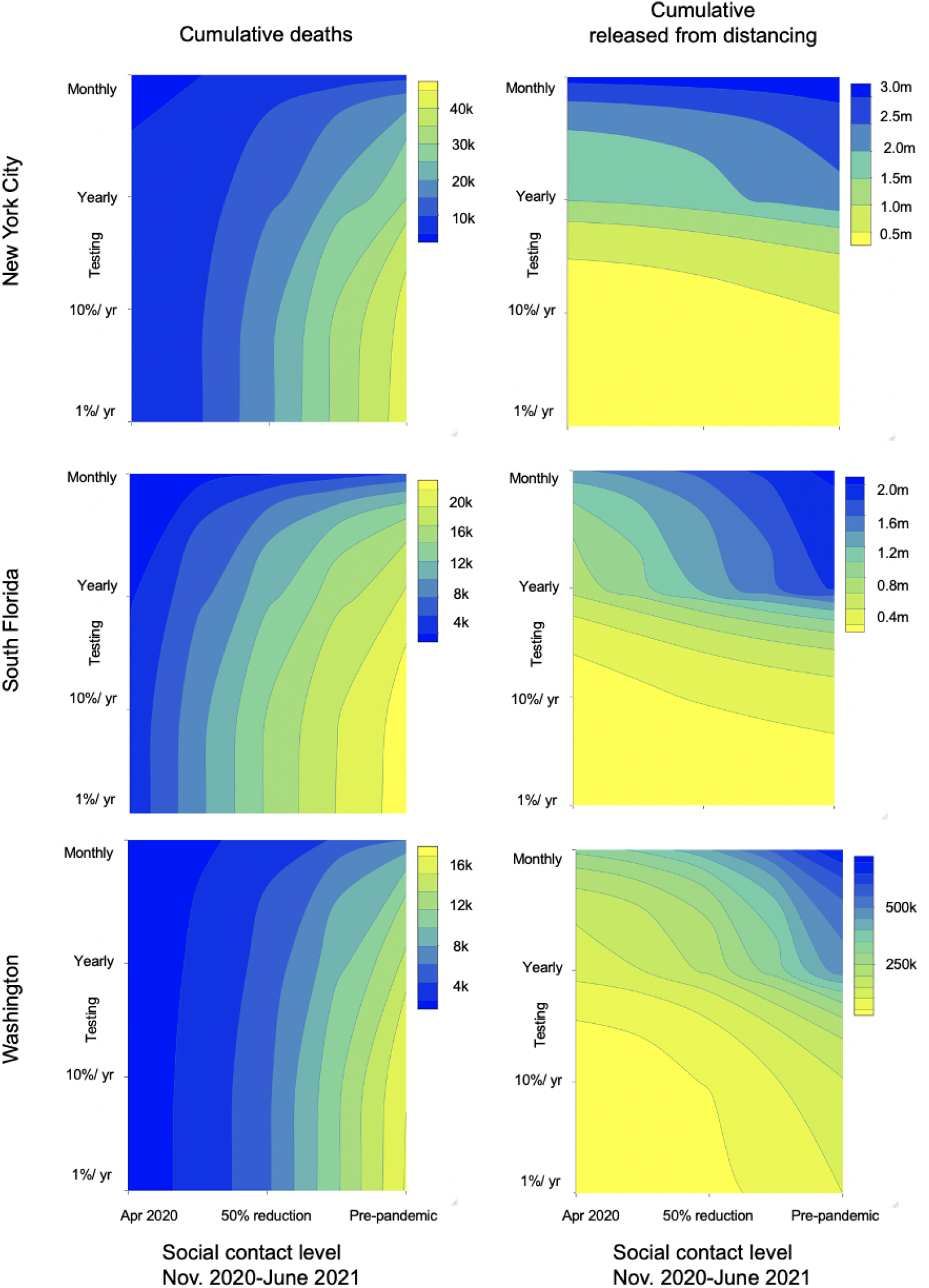
Contour plot of cumulative deaths in each location from November 1, 2020 to June 1, 2021 (left column) and number of people released from social distancing (right column) as a function of the degree of relaxation of social distancing and number of tests per day. The far right of the x-axis corresponds to a pre-pandemic level of contact and the far left corresponds to the contact levels in each location during stay-at-home orders in March-June. The reduction in contact during stay-at-home orders was fitted for each location and is shown by the vertical red dotted line. Both panels assume a test specificity of 99.8% and a shielding factor of 5:1.

### Impacts of serological testing performance on epidemic outcomes and release from social distancing

The value and safety of a serological testing strategy depends on the level of shielding and test specificity. Thus far, our results centered on dynamics enabled by a high-performance test with a specificity of 99.8%, consistent with the high end of the range of reported specificity of available antibody tests, and a suboptimal test with 96% specificity, consistent with the low end of this range (16). If monthly testing with a suboptimal assay (96% specificity) were implemented without shielding, 58-72% of the population would be released from social distancing (72% in New York City, 95% CrI: 46-92%; 58% in South Florida, 95% CrI: 54-61%; and 60% in Washington, 95% CrI: 43-66%) but 85,000 deaths would be expected, more than if no testing were implemented. Increasing the strength of shielding (5:1) while using a suboptimal assay results in fewer deaths (55,000) with 25-48% of the population released from social distancing (61% in New York, 95% CrI: 46-91%; 51% in South Florida, 95% CrI: 47-53%; 44% in Washington, 95% CrI: 38-47%). Overall, adding shielding to a monthly testing strategy results in 30-55% fewer deaths compared to testing at the same frequency without shielding (30% in New York, 95% CrI: 1-33%; 30% in South Florida, 95% CrI: 24-34%; 55% in Washington, 95% CrI: 39-56%). We also set test specificity to 50% to represent a scenario in which antibodies are not a reliable correlate of immunity (i.e., the test is poor at distinguishing between immune and non-immune individuals). If antibodies are not a reliable correlate of protection (which would be counter to current evidence that shows neutralizing antibodies persist for months (25)) then serological testing could lead to more deaths than if not used at all (Figure 4, top panel). We conclude that even if a moderately specific test (96%) is used and antibodies provide a reasonably good correlate of protection, increases in serological testing provide benefits in terms of deaths averted when used with any level of social distancing. (Figure 5)

## Discussion

Strategies are needed to permit safely easing social distancing measures and returning to productive levels of economic and social activity. Our results suggest that widespread serological testing can make a substantial contribution to these efforts. As schools reopen this fall and social distancing measures relax, there is a window of opportunity to reduce the intensity of the pandemic through the use of serological testing, without resorting to further stay-at-home measures or ‘lockdowns.’ In order to explore the impact of this concept in metropolitan areas, we find that maintaining moderate social distancing at the current level of intensity together with widespread serological testing (monthly testing) could relieve 16-35% of local populations from social distancing by June 2021. Moreover, if moderate shielding is employed, a strategy with serological testing results in up to 21,000 fewer deaths than a strategy without testing across the three focal areas. Moderate social distancing and shielding alongside monthly testing results in a flattened curve that provides time to improve treatments and expand healthcare capacity and perhaps roll-out vaccination, which could further reduce mortality rates (26). Thus, serological testing would allow a substantial fraction of the population to return to work and other activities with relative safety, compared to a universal rollback of social distancing policies and without the social and economic costs of a new wave of stay-at-home measures or ‘lockdowns.’

The eventual number of deaths remains uncertain, reflected in the wide, overlapping credible intervals for some testing scenarios. However, sufficiently frequent testing using high-performance tests combined with serological shielding consistently decreased deaths and was consistently able to allow relax social distancing for a substantial fraction of the population. The largest source of uncertainty for estimating future deaths is the extent to which individuals will continue to practice social distancing over the coming months. The width of our credible intervals thus reflects the importance of ongoing social distancing to determining both the trajectory of the U.S. epidemic and the potential impact of any other control interventions. Social distancing was broadly adopted in the initial response (27), but there is a pressing need to continue collecting mobility and mixing data as the epidemic unfolds to better predict risk.

An aggressive testing approach (monthly testing of the whole population) may appear unprecedented but, we argue, is feasible and warranted considering the social and economic impact of the epidemic. Other countries including Germany have implemented serological testing, including repeat testing of the same individuals on a regular (i.e. monthly) basis (28). Implementation in the U.S. would require a significant and rapid scale-up of serological testing capacity. Parallel gains have already been achieved for diagnostic PCR testing; the U.S. expanded testing from fewer than 1,000 tests per day in early March to nearly 250,000 tests per day in mid-May and about 1 million per day by September. Moreover, recently developed serological tests are quicker to perform than RT-PCR, with a potential throughput of 300 tests/hour/machine, compared with 94 RT-PCR tests performed every 3 hours (29,30). While to our knowledge South Florida and Washington have not reported their current serological testing capacity, New York City reported performing a peak of 187,000 tests per week in late March (31). The testing rate in March corresponds to a rate between the yearly testing and 10% per year testing scenarios we include in our analysis and could likely be increased with a concerted focus on antibody testing. To scale up testing capacity on a local level, municipalities would need to take advantage of existing infrastructure (testing locations, healthcare personnel to staff locations, etc.). Highly sensitive and specific bloodspot assays than can be self-administered (32) have also been developed (33), and could be used for this purpose, easing some of the logistical issues with widespread testing.

Even if testing can be scaled up, legal and ethical concerns remain. Requiring evidence of a positive test to return to work may create incentives for individuals to misrepresent their immune status or intentionally get infected. A large-scale testing program must consider how such policies might reinforce social disparities and guard against inequities in test availability (34). However, the use of serological testing to reduce new transmission can also be supported on ethical grounds, insofar as serological tests are sufficiently accurate (35). Indeed, current policy approaches to utilize Covid-19 positive, asymptomatic health-care workers to treat hospitalized patients suggests that principled, test-driven policies are needed. Relatedly, a history of a positive antibody test should not change the clinical care of individuals with respiratory symptoms suggestive of SARS-CoV-2 infection if a PCR diagnostic test would otherwise be indicated.

While others have raised concerns that using serological tests to relax social distancing could increase population risk (36), we show that coupling serological testing with available diagnostic tests with immune shielding can form the basis of a successful strategy. If testing employs the most specific assays available, the false positive proportion would remain low and decrease over time as seroprevalence increases. As such, deploying immune individuals such that they are responsible for more interactions than susceptible individuals will reduce risk. If shielding is not employed, this benefit disappears, and testing can become a liability – reinforcing the critical need to combine serological testing with a shielding strategy. In our model, cumulative deaths are not substantially impacted by the false positive rate: deaths are similar under scenarios assuming a sub-optimally specific test (96% specificity) and a high-performance test (99.8% specificity) except at very high testing rates. Thus, false positives are unlikely to substantively impact population-level risk at levels of specificity reported by most authorized serological tests (16,37).

Our models assumed random allocation of serological testing. In practice, targeting testing to specific groups, such as healthcare workers, nursing home care providers, food service employees, or contacts of confirmed or suspected cases might increase efficiency by increasing the test-positive rate (and consequently, cost-effectiveness (38)), allowing for similar numbers of individuals to be released from social distancing at lower testing levels. This strategy would also decrease the false positive rate, an important consideration if a less specific test is used (39). Many healthcare organizations have already begun to offer antibody testing to their employees (40). The use of serological testing and shielding within healthcare settings represents a potential application of a more targeted strategy and may also help to allow ethical concerns on misrepresentation of antibody status and/or intentionally seeking infections.

There remains much to learn about immunity to SARS-CoV-2 and we have made three critical simplifying assumptions in our model. First, we assume that antibodies are immediately detectable after resolution of infection. In reality, this generally occurs between 11 and 14 days post infection (41). A small fraction of recent infections would be undetected, but this would have a minor effect on our results. Second, we assume that immunity lasts for at least a year. Given that the virus has only been circulating for about nine months in humans, both the duration of antibody protection and the extent to which those antibodies protect against future infections remains unclear. However, the great majority individuals who are infected seroconvert (14), and ongoing studies of SARS-CoV-2 show that antibodies persist for at least months (41). Evidence is emerging that antibodies can wane in a matter of months though waning antibodies does not necessarily imply the loss of immune protection (42). Third, we assume that antibodies detected by serology are a correlate of protection. While antibody levels have been shown to wane after several months, especially for individuals with mild infection (25,42), antibody kinetics of SARS-CoV-1 and MERS suggest that protection will last for at least months and as long as several years (43). Reinfections, while documented, have been extremely rare.

While we have focused our analysis on serological testing, using the principles of serological shielding to reduce risks of infection for susceptible individuals also applies to other public health interventions (44). As of October 2020, many phase III trials of vaccines against SARS-CoV-2 are underway and efficacy readouts are expected at the end of 2020 or early 2021. If and when an effective vaccine becomes available, vaccinated individuals could also be preferentially placed in high contact positions to serve as immune ‘shields.’ This could both allow vaccinated individuals to resume more normal levels of social and economic activity and also allow herd immunity to be reached more quickly. However, even after a vaccine becomes licensed, scaling up access to vaccination will likely take several months, with individuals at highest risk likely receiving vaccines first (45). In the interim, using serological testing as an additional strategy to reduce transmission could be helpful to reduce the public health and socioeconomic impacts of the pandemic.

A serological testing strategy is one component of the public health response to COVID-19, alongside viral testing, isolation and rigorous contact tracing. If implemented simultaneously, such measures would likely reduce the extent of shielding required to achieve the same benefit, in addition to further reducing overall transmission. In summary, our results suggest a role for large-scale serological testing programs in the public health response to COVID-19. While maintaining a degree of social distancing, serology can be used to allow people with positive test results to return to work and other activities while mitigating the health impacts of COVID-19.

## Materials and Methods

### Model

We modeled the transmission dynamics of SARS-CoV-2 using a deterministic, compartmental SEIR-like model. (Figure 1) We assume that after a latent period, infected individuals progress to either asymptomatic or symptomatic infection. A fraction of symptomatic cases are hospitalized, with a subset of those requiring critical care. Surviving cases, both asymptomatic and symptomatic, recover and are assumed to be immune to reinfection. All individuals who have not tested positive and are not currently experiencing symptoms of respiratory illness are eligible to be tested and all hospitalized cases are tested prior to discharge. Recovered individuals are moved to the test-positive group at a rate that is a function of test sensitivity. Susceptible, latently infected, and asymptomatic cases may falsely test positive and are moved to the test-positive group at a rate that is a function of test specificity. False positives may become infected, but the inaccuracy of their test result is not recognized unless they develop symptoms that are sufficiently severe to warrant hospitalization and health providers correctly diagnose COVID-19, overriding the history of a positive antibody test. The ordinary differential equations corresponding to this model are included in section S1 of the SI. All models were run in R version 3.6.2 using the package ‘deSolve’. Code is available at https://github.com/lopmanlab/Serological_Shielding.

There are three age groups represented in the model: children and young adults (<20 years), working adults (20-64 years), and elderly (65+ years). We modeled age-specific mixing based on POLYMOD data adapted to the population structure in the United States (46,47). Contacts in this survey were reported based on whether they occurred at home, school, work, or another location. All baseline social contact matrices were based on Prem et *al*. (47) and calculated to be symmetric using the *socialmixr* R package.

‘General’ social distancing began on the day that stay-at-home orders were enacted in each location. Although adherence to these measures varied and is generally difficult to measure, we made several assumptions about how these policies changed location-specific contacts. First, we assume that under these measures, all contacts at school and daycare were eliminated and that contacts outside of home, work, and school (‘other’) locations were reduced by a fraction, which was fitted for each metropolitan location. We assume that contacts at home remained unchanged. To address differences in work-based contacts by occupation types, we classified the working adult population into three subgroups based on occupation: (i) those with occupations that enable them to work exclusively from home during social distancing, (ii) those continuing to work but reduced their contacts at work (e.g., customer-facing occupations such as retail), and (iii) those continuing to work with no change in their contact patterns (e.g., frontline healthcare workers). The percent reduction in ‘other’ contacts and percent contact reductions at work for essential workers who could reduce their contacts was fitted (see next section).

This period of intense social distancing lasts until stay-at-home orders are lifted (referred to as ‘reopening’) in each location. After this date, we assume that schools and daycares remain closed but that social distancing measures for the general population can be relaxed, by allowing work and other contacts to be increased. To represent this, we scale contacts at work and ‘other’ locations to a proportion of their value under general social distancing based on a scalar constant, c, such that c = 1 is equivalent to the scenario in which social distancing measures as put into place in March are maintained and c = 0 is equivalent to a return to pre-pandemic contact levels for both ‘work’ and ‘other’ contacts for essential workers and pre-pandemic contact levels for ‘other’ contacts for all other groups. Based on local policies, we assume that children returned to school and daycare on September 1, 2020 in South Florida and October 1, 2020 in Washington and New York City. To account for the fact that schools have taken a variety of measures to reduce contact among students, we assumed that children are halving (50%) their pre-pandemic contacts at school.

### Fitting data

We fit the model for each location to deaths reported due to COVID-19 from March to July 2020 (20) as well as seroprevalence data (12) using a Markov Chain Monte Carlo (MCMC) approach using the MCMCstat toolbox (https://mjlaine.github.io/mcmcstat/). Each location was defined using the same counties as were included in a CDC-led seroprevalence study (12). For each location, we estimated the six parameters listed in Table S1. We ran ten, randomly seeded chains of 100,000 iterations each (95,000 burn-in; 5,000 samples). To infer the initial conditions, we first calculated the number of weeks between the first reported death in each location and the first week where the cumulative death toll exceeded 10. Using region-specific conditions (population demographics, stay-at-home order enactment and lifting dates, and death data), we initialize an epidemic consisting of a single exposed adult, forward simulated until a death count threshold was met, and then the population distribution was used as the initial condition for subsequent intervention scenarios.

We used a Poisson likelihood function that included penalty terms for *R*_*0*_ (expected *R*_*0*_ = 3.0), the cumulative midpoint and final number of deaths in each location, as well as population-level seroprevalence estimated for each location from Havers *et al*. (12) We checked for chain convergence using the Gelman-Rubin diagnostic (Figure S1). Figures S2, S5, and S8 show the trace plots for each model, and Figures S3, S6, and S9 show the resulting joint distributions of estimated parameters. The consistency between the fitted model and death/seroprevalence data for each location is shown in Figures S4, S7, and S10. After fitting to death data spanning March to July 2020, we use fitted parameters to forward-simulate the epidemic through June 1 in each location.

To capture uncertainty in model predictions, we randomly sampled 20 parameter sets in the final 5,000 iterations in each chain for each location. For Washington, one of the chains did not converge, so we used sampled parameter sets from the other 9 chains. For South Florida and New York, all 10 chains converged to the same final state, so we randomly sampled parameter sets from all 10 chains. This resulted in 200 randomly sampled parameter sets for New York and South Florida and 180 for Washington. We simulated the epidemic forward using each parameter set for the key testing and shielding interventions we report in the text. We report the middle 95% of the distribution of outcomes from these runs as our credible intervals. We have uploaded a supplementary .csv file with the number of deaths, critical care cases, cumulative incidence, and the fraction of the population released from social distancing after one year from each of these simulations as supplemental material.

## Data Availability

Transmission model code and incidence data used are available at the links provided

https://coronavirus.jhu.edu/map.html

https://github.com/lopmanlab/Serological_Shielding

https://usafacts.org/visualizations/coronavirus-covid-19-spread-map/

## Acknowledgments

We thank Timothy Lash, Andreas Handel, Carly Adams, Julia Baker, Carol Liu and Avnika Amin for useful comments.

## Author contributions

The model was designed by ANM and KNN, extended from an earlier version by JSW and CYZ. All authors designed model simulations, ANM, KNN, and CYZ conducted the analysis with input from DD, BAL, and JSW; DD and CYZ led the model-fitting and ANM, KNN, and BAL wrote the first draft of the manuscript. All authors contributed to editing the manuscript. JSW, DD, and CYZ provided critical review of the code, results, and conclusions. ANM, KNN, JSW, and BAL designed the study.

## Data and materials availability

Data used for model calibration are available from ‘https://github.com/nytimes/covid-19-data’ and ‘https://covidtracking.com/api’. Code is available at ‘https://github.com/lopmanlab/Serological_Shielding’

