## Supplemental material for "Modeling serological testing to inform relaxation of social distancing for COVID-19 control"

#### S1. Model Equations

The ordinary differential equations describing the model are shown below for group i. The positive test group is denoted with the '+' sign.

[a]These authors contributed equally to this work
[b]These authors contributed equally to this work

$$
\begin{aligned}
\frac{dS_i}{dt} &= -\lambda_i(t)S_i - (1-sp)\times test_i(t)S_i \\
\frac{dE_i}{dt} &= \lambda_i(t)S_i - \gamma_e E_i - (1-sp)\times test_i(t)E_i \\
\frac{dI_{s,i}}{dt} &= \gamma_e E_i p - \gamma_s I_{s,i} \\
\frac{dI_{a,i}}{dt} &= \gamma_e E_i(1-p) - \gamma_a I_{a,i} - (1-sp)\times test_i(t)I_{a,i} \\
\frac{dH_{s,i}}{dt} &= \gamma_s I_{s,i}(Hosp_i - Crit_i) + \gamma_s I^+_{s,i}(Hosp_i - Crit_i) - \gamma_{hs}H_{s,i} \\
\frac{dH_{c,i}}{dt} &= \gamma_s I_{s,i}Crit_i + \gamma_s I^+_{s,i}Crit_i - \gamma_{hc}H_{c,i} \\
\frac{dD_i}{dt} &= \gamma_{hc}H_{c,i}Die_i \\
\frac{dR_i}{dt} &= (1-Hosp_i)\gamma_s I_{s,i} + \gamma_a I_{a,i} + (1-se)\times m_2(t)\gamma_{hs}H_{s,i} + (1-se)\times m_2(t)\gamma_{hc}H_{c,i}(1-Die_i) \\
&\quad - se\times test_i(t)R_i + m_1(t)\gamma_{hs}H_{s,i} + m_1(t)\gamma_{hc}H_{c,i}(1-Die_i) \\
\frac{dS^+_i}{dt} &= (1-sp)test_i(t)S_i - \lambda^+_i(t)S^+_i \\
\frac{dE^+_i}{dt} &= (1-sp)test_i(t)E_i + \lambda^+_i(t)S^+_i - \gamma_e E^+_i \\
\frac{dI^+_{s,i}}{dt} &= \gamma_e E^+_i p - \gamma_s I^+_{s,i} \\
\frac{dI^+_{a,i}}{dt} &= (1-sp)\times test_i(t)I_{a,i} + \gamma_e E^+_i(1-p) - \gamma_a I^+_{a,i} \\
\frac{dR^+_i}{dt} &= se\times test_i(t)R_i + se\gamma_{hc}m_2(t)H^+_{c,i}(1-Die_i) + \\
&\quad se\times m_2(t)\gamma_{hs}H_{s,i} + (1-Hosp_i)\gamma_s I^+_{s,i} + \gamma_a I^+_{a,i}
\end{aligned}
$$

$m_1(t)$ and $m_2(t)$ are defined as follows ($t_{start}$ is when testing starts, on November 1, 2020):

$$
m_1(t) = \begin{cases} 1 & t < t_{start} \\ 0 & t \geq t_{start} \end{cases}
$$

$$
m_2(t) = \begin{cases} 0 & t < t_{start} \\ 1 & t \geq t_{start} \end{cases}
$$

The force of infection $\lambda(t)$ for the $i$-th group is a function of the number of social contacts for age group $i$ with each subgroup $j$ at time $t$ ($x_{i,j}(t)$), the probability of infection given contact ($q$), the number of infections in each group at time t ($infec_j(t)$), and the population size of each group at time $t$ ($n_j(t)$). The overall equation for $\lambda_i(t)$ is shown below:

$$\lambda_i(t) = q\Bigg[\frac{x_{i,ch}(t)infect_{ch}(t)}{n_{ch}(t)} + \frac{x_{i,ch^+}(t)infect_{ch^+}(t)}{n_{ch^+}(t)} + \frac{x_{i,ad}(t)infect_{ad}(t)}{n_{ad}(t)}) + \frac{x_{i,ad^+}(t)infect_{ad^+}(t)}{n_{ad^+}(t)} + \frac{x_{i,rc}(t)infect_{rc}(t)}{n_{rc}(t)} + \frac{x_{i,rc^+}(t)infect_{rc^+}(t)}{n_{rc^+}(t)} + \frac{x_{i,fc}(t)infect_{fc}(t)}{n_{fc}(t)}) + \frac{x_{i,fc^+}(t)infect_{fc^+}(t)}{n_{fc^+}(t)} + \frac{x_{i,el}(t)infect_{el}(t)}{n_{el}(t)}) + \frac{x_{i,el^+}(t)infect_{el^+}(t)}{n_{el^+}(t)}\Bigg]$$

$i$ and $j$ take values of $ch$ (children age 0-19 years), $ad$ (adults age 20-65 years who are not working or working from home), $el$ (older adults 65+ years of age), $rc$ (adults at work in 'reduced-contact' occupations, where they have fewer contacts than pre-pandemic), $fc$ (adults as work in full contact occupations, where they have the same number of contacts as pre-pandemic).

The number of infectious individuals by age group and test status is equal to the sum of documented (symptomatic) cases and a fraction of the undocumented (asymptomatic) cases, where this fraction $asy$ corresponds to the relative infectiousness of undocumented cases. This is shown below for children:

$$infect_{ch}(t) = asyI_{a,ch} + I_{s,ch}$$

### S2. Model parameters

We defined the three metropolitan areas in the same way as Havers et al [1]. For Washington Puget Sound metropolitan region, we included death data from King, Snohomish, Pierce, Kitsap and Grays Harbor counties. For the New York City metropolitan region, we included data from Manhattan, Bronx, Queens, Kings, and Nassau Counties. For the South Florida metropolitan region, we included data from Miami-Dade, Broward, Palm Beach, and Martin counties. These same counties were also used to derive age-specific population sizes for each region.

Parameters used in the model simulations are shown in Table S1. We assume that the size of the working population is stable over the duration of the simulation. Although this may not be the case as unemployment increases throughout the pandemic, the rate at which unemployment has changed so far has been time-varying and its future trajectory is unknown.

Where possible, parameters were taken from prior literature. However, data from the initial stages of the outbreak were fitted for the probability of infection per contact ($q$), the fraction of infections symptomatic ($p$), the initial intensity of social distancing for workplace contacts ($p_{reduced}$) and for other contacts ($sd_{other}$). These parameters were fitted to initial outbreak dynamics from each of the three metropolitan locations. While $q$ does not vary with time, we acknowledge that some control measures such as masking may result in changes in the probability of infection given contact over the course of the pandemic. In our model, changes in the contact matrix based on both the initial strength of social distancing ($p_{reduced}$ and $sd_{other}$) and its strength after reopening captures the contribution of both reducing the number of social contacts and changes in the likelihood of transmission from those contacts based on mask use and physical distancing.

We used the dates that stay-at-home orders were enacted, and later lifted, in each location and the dates corresponding to when local schools opened for at least partial in-person instruction in each location to inform dates of reopening in the model.

### S3. Model fitting

#### Initial Conditions

We first calculate the number of weeks between the first death and the first week where the cumulative death toll exceeds 10. For example, in the South Florida region, the first death was reported the week of March 18, 2020. The two subsequent weeks saw the death toll rise to 6 and 42. Using region-specific conditions (population demographics, stay-at-home order and lift dates, and deaths data), we initialize an epidemic consisting of a single exposed adult ($a0$). We use baseline estimates for the parameters we aim to fit later ($q$ = 0.0451, $c$ = 1, $symptomatic\_fraction = 0.14$, $sd_{other} = 0.25$, $p_{reduced} = 0.1$). Note that $symptomatic\_fraction$ is equivalent to p in Table 2. We forward simulate a single-origin epidemic until the modeled number of deaths exceeds the number of deaths reported in the second week after the first death (in South Florida, 42 deaths), then use the distribution two weeks prior as our initial conditions. For MCMC fits, we additionally allow an error term, $init_{scale}$, such that the calculated distribution was scaled by $(1 + init_{scale})$ for each chain iteration. For fitting, we constrained $q$ to $[0, 0.07]$ and $init_{scale}$ to $[0, \inf)$. All other parameters were constrained to $[0, 1]$.

| Parameter | Code | Value | Units | Source(s) |
|---|---|---|---|---|
| **Natural history** | | | | |
| Latent period | $\gamma_e$ | 1/3 | 1/days | [2] |
| Recovery rate, asymptomatic infections | $\gamma_a$ | 1/7 | 1/days | [3] |
| Recovery rate, symptomatic infections | $\gamma_s$ | 1/7 | 1/days | [3] |
| Recovery rate, hospitalized cases | $\gamma_{hs}$ | 1/5 | 1/days | [4] |
| Recovery rate, critical care cases | $\gamma_{hc}$ | 1/7 | 1/days | [4] |
| Relative infectiousness of asymptomatic infections | $asy$ | 0.55 | – | [5, 6] |
| **Hospitalization** | | | | |
| Probability of hospitalization | | | | [7] |
| Children | $Hosp_{ch}$ | 0.061 | – | |
| Adults | $Hosp_{ad}$ | 0.182 | – | |
| Elderly | $Hosp_{el}$ | 0.417 | – | |
| Probability of requiring critical care | | | | [7] |
| Children | $Crit_{ch}$ | 0 | – | |
| Adults | $Critc_{ad}$ | 0.063 | – | |
| Elderly | $Crit_{el}$ | 0.173 | – | |
| Probability of death among critical care patients | | | | |
| Children | $Die_{ch}$ | 0 | – | [8] |
| Adults | $Die_{ad}$ | 0.5 | – | [5] |
| Elderly | $Die_{el}$ | 0.5 | – | [5] |
| **Test features** | | | | |
| Sensitivity | $se$ | 1.00 | – | [9] |
| Specificity | $sp$ | 0.998 (0.5, 0.998) | – | [9] |
| **Population features** | | | | |
| Population size | | | | [10] |
| New York City | | 9.43e6 | people | |
| South Florida | | 6.17e6 | people | |
| Washington Puget Sound | | 4.06e6 | people | |
| Baseline contact rates by age | $x_{i,j}$ | see matrices | | [11] |
| Adult working population segments, size | | | | [12] |
| Exclusive work from home occupations | $n_{home}$ | $0.316 \times n_{adult}$ | people | |
| Reduced contact occupations | $n_{reduced}$ | $0.628 \times n_{adult}$ | people | |
| Full contact occupations | $n_{full}$ | $0.057 \times n_{adult}$ | people | |
| Fraction of adult population (20-65 years) in workforce | | 1.0 | | assumption |
| **Intervention parameters** | | | | |
| Shielding | $\alpha$ | (0, 9) | per contact | varied |
| Fraction tested | $test_i(t)$ | (0, 0.03) | proportion per day | varied |

Table S1: Fixed parameters across locations used in model simulations. Values shown in parentheses represent a range, used to perform sensitivity analysis.

| Parameter | Code | New York | South Florida | Washington | Units | Source(s) |
|---|---|---|---|---|---|---|
| **Fitted parameters** | | | | | | |
| Fraction symptomatic[1] | $p$ [2] | 0.480 | 0.203 | 0.475 | – | fitted |
| | | (0.407, 0.555) | (0.185, 0.224) | (0.497, 0.519) | | |
| Probability of infection per contact[3] | $q$ | 0.034 | 0.043 | 0.023 | 1/contact | fitted |
| | | (0.032, 0.036) | (0.039, 0.045) | (0.022, 0.022) | | |
| | | (1.91, 2.12) | (2.57, 2.67) | (1.28, 1.31) | | |
| Strength of social distancing maintained[1] | $c$ | 0.491 | 0.974 | 0.126 | – | fitted |
| | | (0.026, 0.965) | (0.923, 0.999) | (0.009, 0.337) | | |
| Fraction of work contacts maintained ($rc$ workers)[1] | $p.reduced$ | 0.323 | 0.367 | 0.800 ) | – | fitted |
| | | (0.022, 0.678) | (0.016, 0.776) | (0.545, 0.979 | | |
| for $rc$ occupations | | | | | | |
| Fraction of 'other' contacts maintained[1] | $sd.other$ [5] | 0.106 | 0.195 | 0.168 | – | fitted |
| | | (0.007, 0.555) | (0.014, 0.398) | (0.056, 0.294) | | |
| Initial conditions scaling term[4] | – | | | | – | fitted |
| | | (0.005, 0.446) | (0.004, 0.490) | (0.0004, 0.042) | | |
| Epidemic start date | $t_0$ | Feb 20 | Feb 27 | Feb 13 | date | fitted |
| Basic reproduction number | $R_0$ | 2.04 | 2.63 | 1.33 | – | calculated from fitted value |
| **Population features (fixed)** | | | | | | |
| Baseline contact rates by age | $x_{i,j}$ | | see matrices | | | [11] |
| US population age groups, size | | | | | | [13, 10] |
| Children | $n_{child}$ | 2208909 | 1398572 | 974924 | people | |
| Adults | $n_{adult}$ | 5944919 | 3679331 | 2563244 | people | |
| Elderly | $n_{elderly}$ | 1283369 | 1090711 | 526665 | people | |
| **Intervention timing (fixed)** | | | | | | |
| Stay at home order start | – | Mar 16 | Mar 17 | Mar 16 | date | [14, 15, 16] |
| Reopening | – | June 8 | May 20 | May 31 | date | [14, 15, 16] |
| School reopening | – | Oct 1 | Sept 1 | Oct 1 | date | assumption |

1 Constrained so fitted value would fall between 0 and 1

2 Note that $p$ is equivalent to $symptomatic\_fraction$ in Figures S1-S9

3 Constrained so fitted value would fall between 0 and 0.07

4 Constrained so fitted value would fall between 0 and infinity

5 Note that $sd.other$ is equivalent to $socialDistancing\_other$ in Figures S1-S9

Table S2: Location-specific parameters used in model simulations. Values shown for fitted parameters are the maximum likelihood estimate, used for main text simulations in Figures 2 and 3, and values in parentheses represent the 95% CI, used to calculate simulation intervals

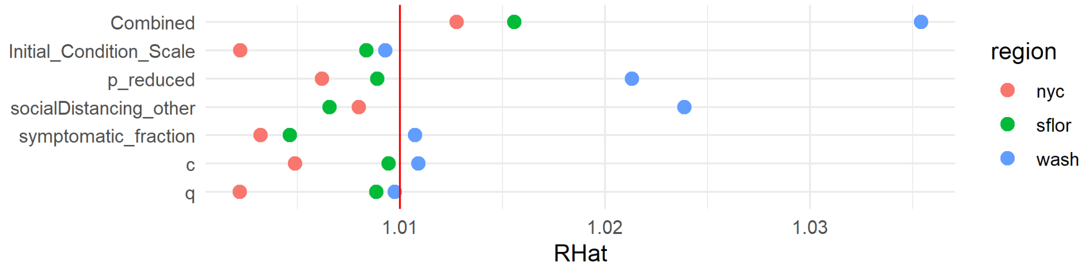


Figure S1: Gelman-Rubin diagnostics plot. We calculate Gelman-Rubin $\widehat{R}$ values for chain convergence among all 10 chains for New York City and South Florida. We excluded chain 3 for Washington because it did not converge. $\widehat{R}$ values for all parameters were below 1.1, with the majority under the 1.01 convergence threshold. [17]

#### MCMC

Using Markov Chain Monte Carlo (MCMC) we estimate the six model parameters listed in Table S2. We checked for chain convergence using the Gelman-Rubin diagnostic (Figure S1). Figures S2, S5, and S8 show the resulting trace plots, and Figures S3, S6, and S9 show the resulting joint distributions.

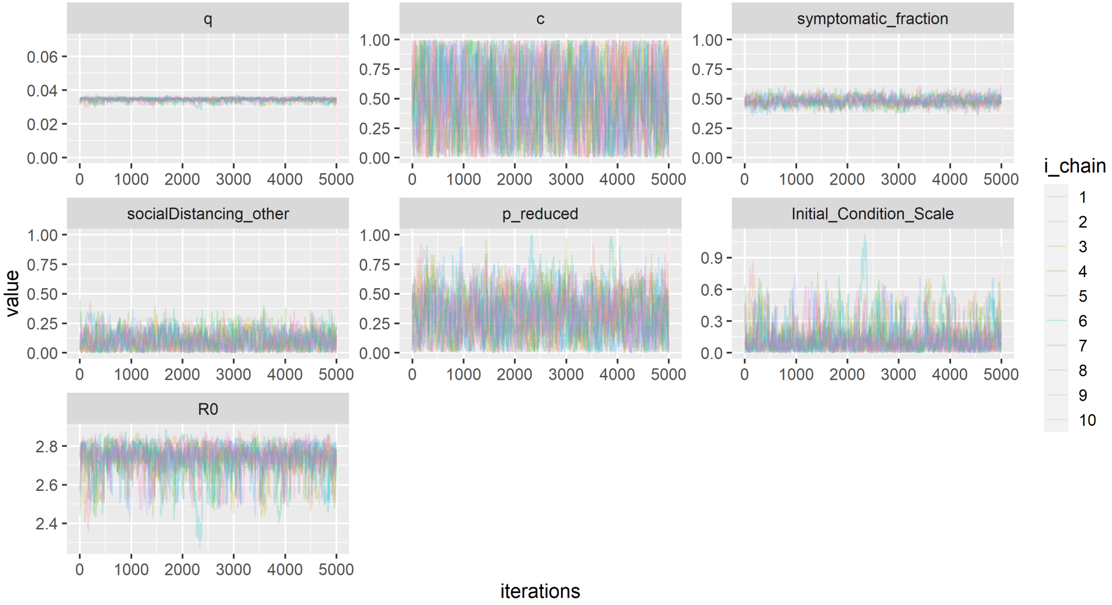


Figure S2: New York City Post-burn-in MCMC Traceplots. Y-axis ranges are limited to show parameter search space constraints. $R_0$ traces are calculated from $q$ and $symptomatic\_fraction$ traces.

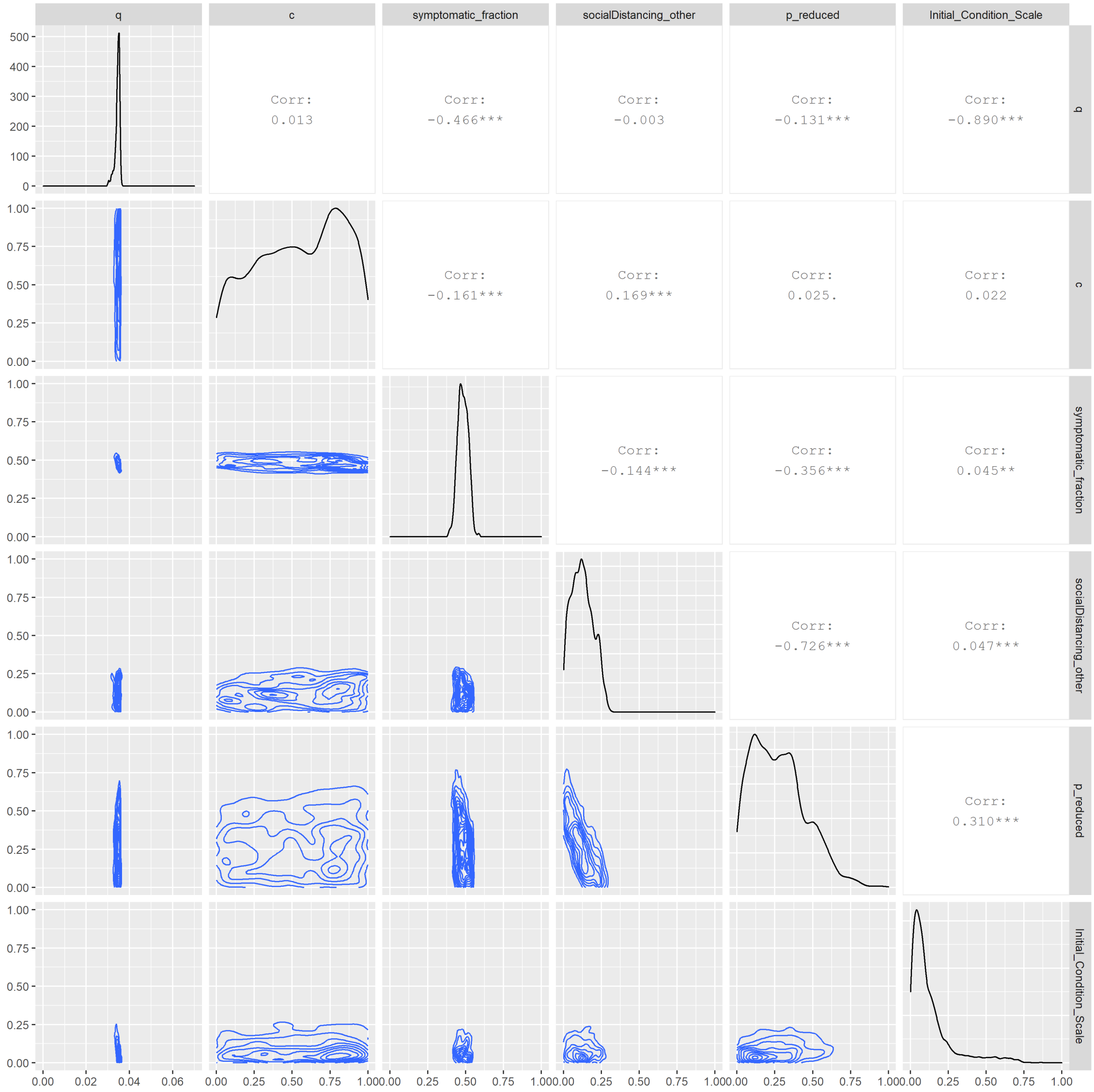


Figure S3: New York City Pairwise Parameter Correlations. Joint posterior distributions for pairs of parameters fitted to data from the New York City region for weekly reported deaths and seroprevalence estimates. Represented is the chain initialized with a minsearch algorithm.

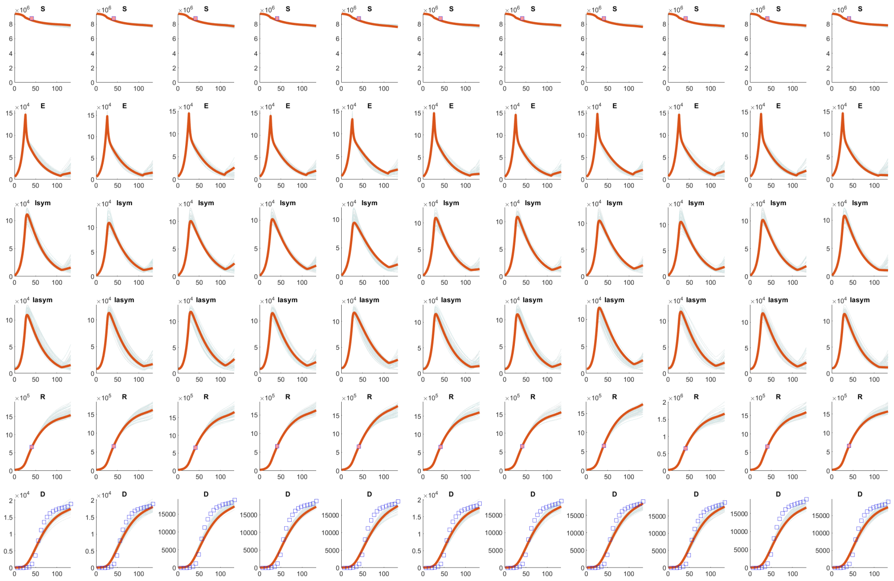

Figure S4: New York City Model Fits. We forward simulate a model using the maximum likelihood estimates of each parameter (red) and 100 random parameter set draws from the posterior distribution (gray). Cumulative deaths are plotted (blue squares), in addition to seroprevalence estimates (orange squares) in each location. Each column corresponds to an MCMC chain, with the first column corresponds to a chain which was initially seeded using a constrained 'minsearch' algorithm. Columns 2-11 correspond to MCMC chains 1-10, which are seeded randomly.

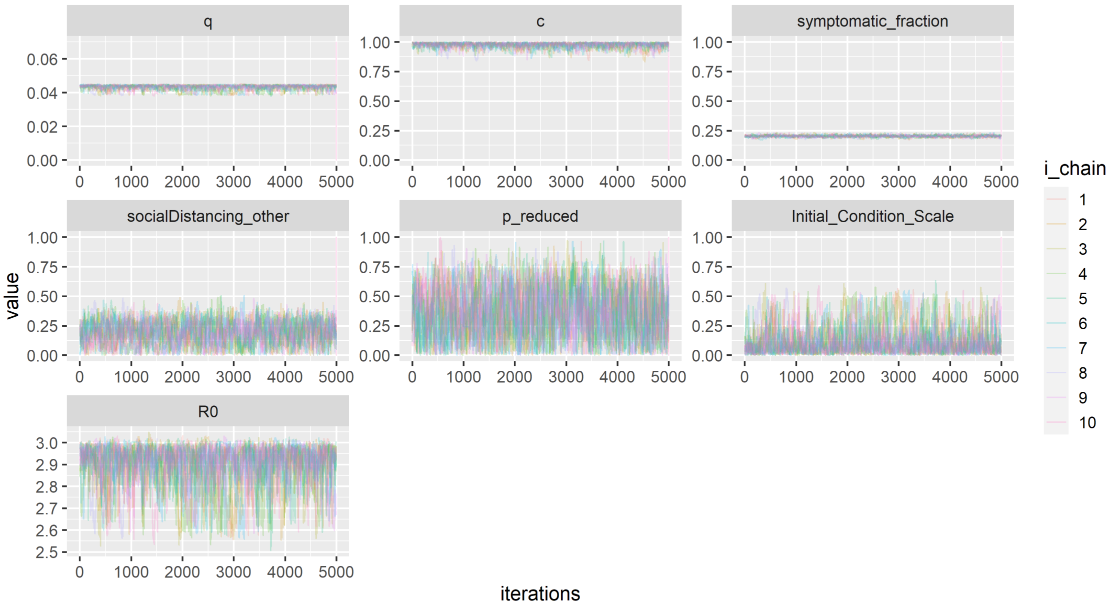


Figure S5: South Florida Post-burn-in MCMC Traceplots. Y-axis ranges are limited to show parameter search space constraints. $R_0$ traces are calculated from $q$ and $symptomatic\_fraction$ traces.

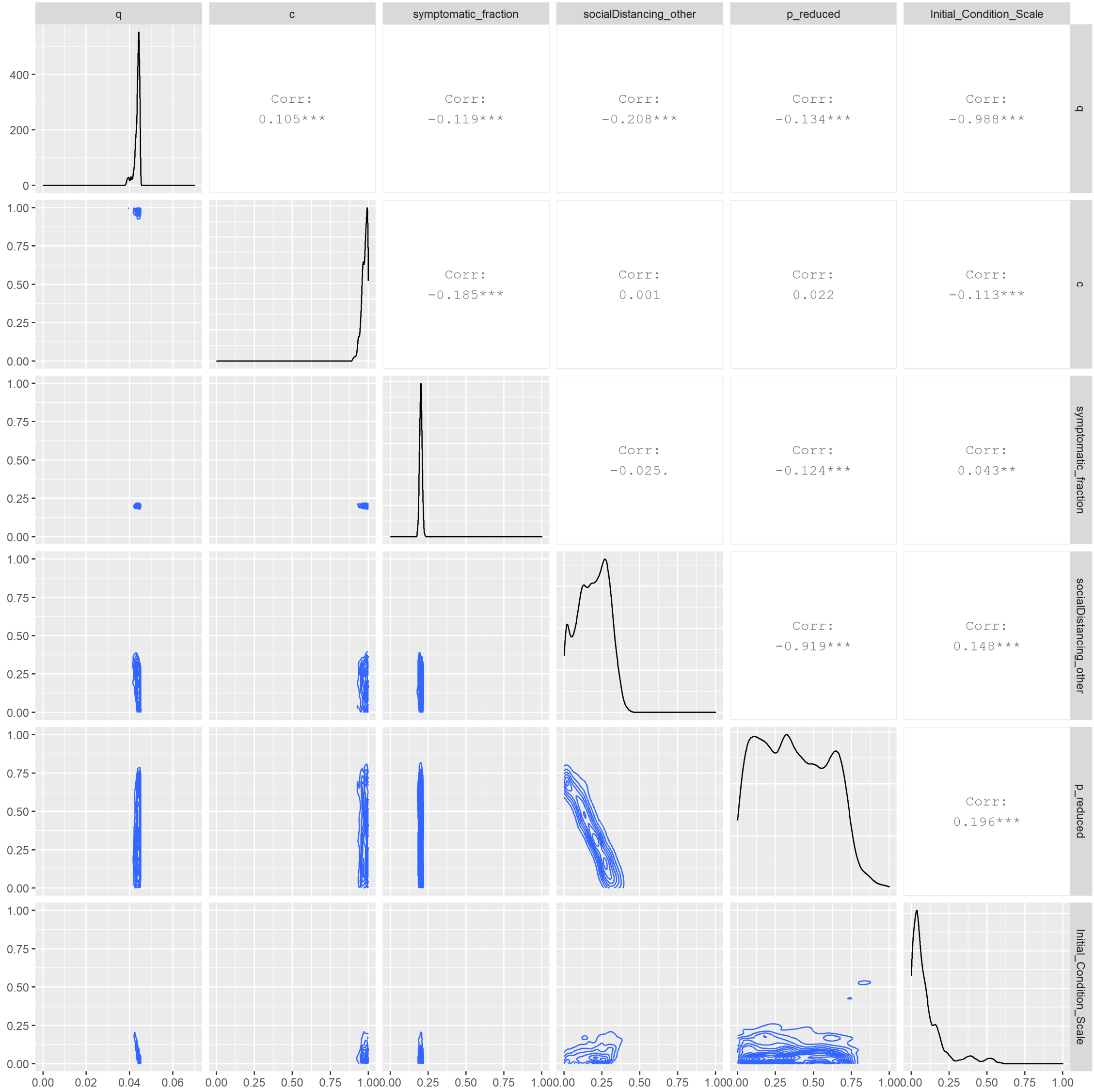


Figure S6: South Florida Pairwise Parameter Correlations. Joint posterior distributions for pairs of parameters fitted to data from the New York City region for weekly reported deaths and seroprevalence estimates.

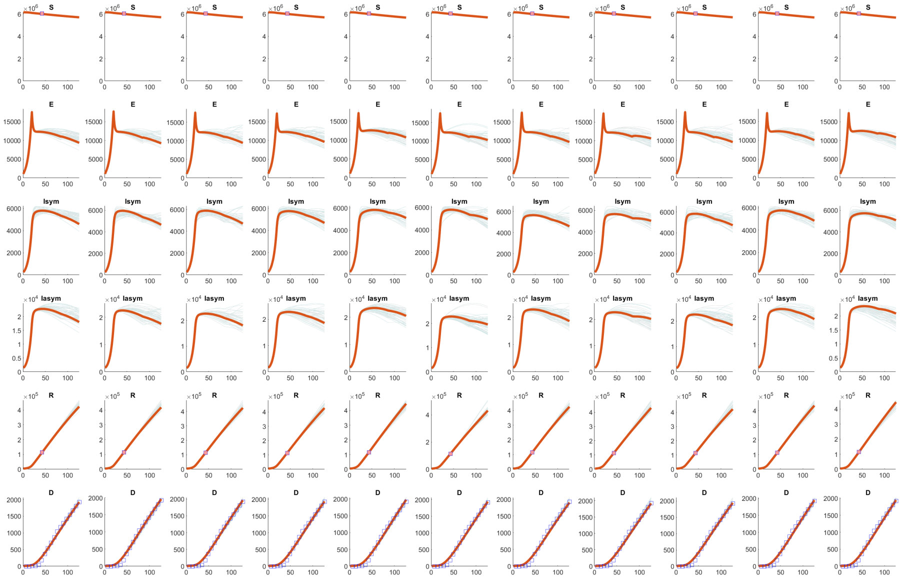


Figure S7: South Florida Model Fits. We forward simulate a model using the maximum likelihood estimates of each parameter (red) and 100 random parameter set draws from the posterior distribution (gray). Cumulative deaths are plotted (blue squares), in addition to seroprevalence estimates (orange squares) in each location. Each column corresponds to an MCMC chain, with the first column corresponds to a chain which was initially seeded using a constrained 'minsearch' algorithm. Columns 2-11 correspond to MCMC chains 1-10, which are seeded randomly.

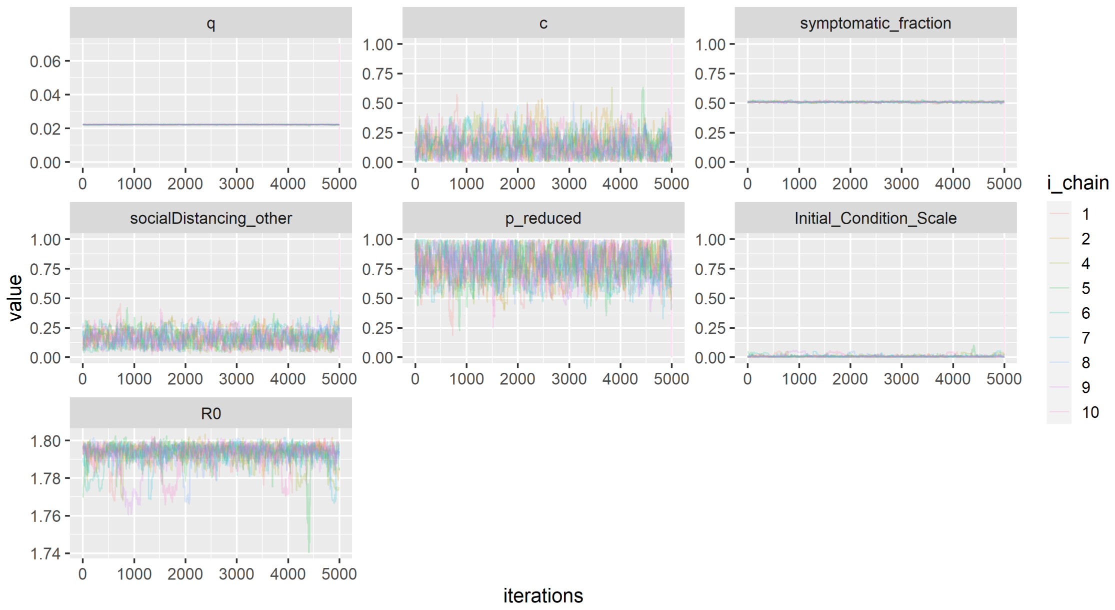


Figure S8: Washington Puget Sound Post-burn-in MCMC Traceplots. Y-axis ranges are limited to show parameter search space constraints. $R_0$ traces are calculated from $q$ and $symptomatic\_fraction$ traces. Note that chain 3, which did not converge, was excluded from analyses.

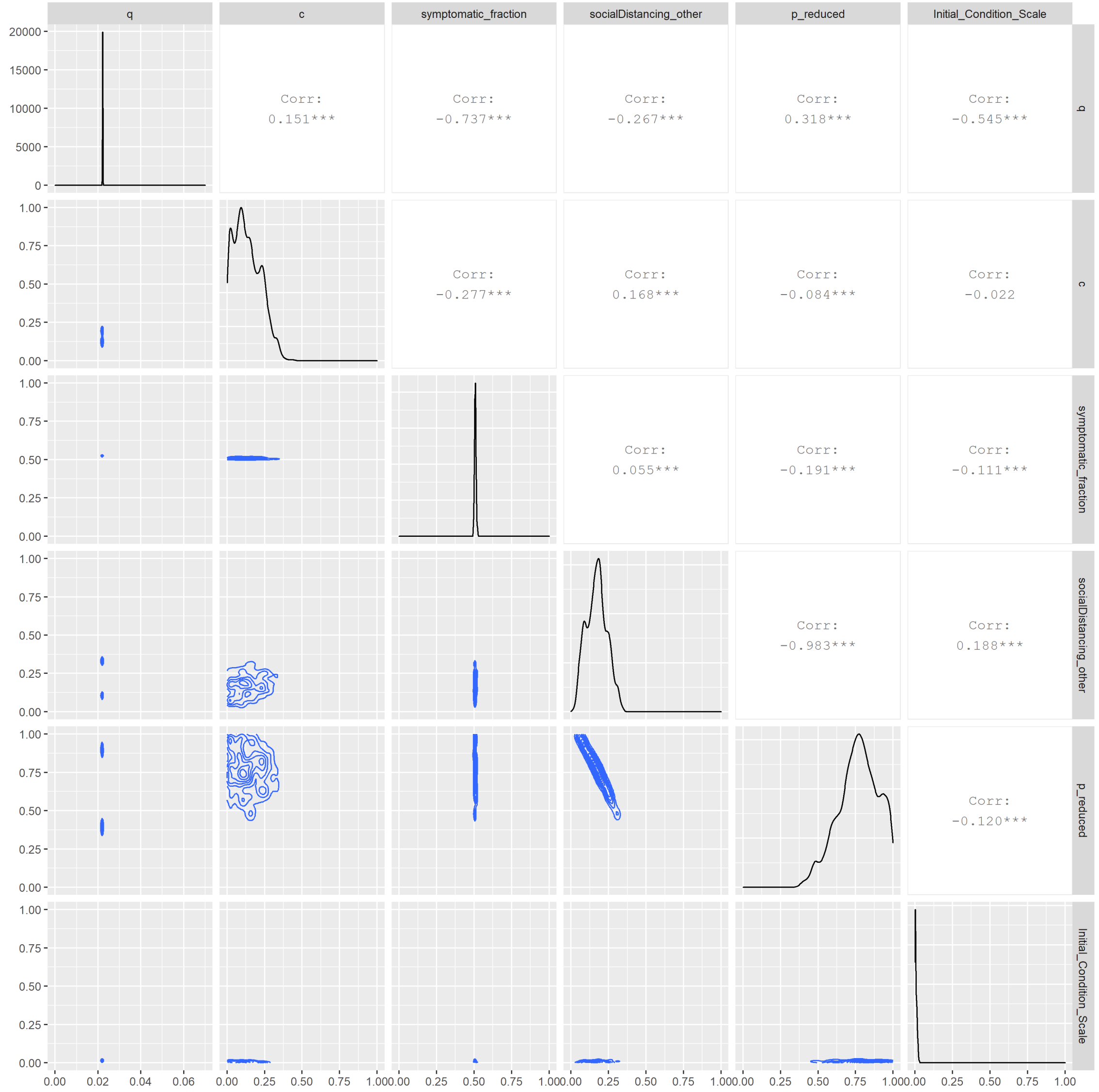


Figure S9: Washington Pairwise Parameter Correlations. Joint posterior distributions for pairs of parameters fitted to data from the New York City region for weekly reported deaths and seroprevalence estimates. Represented is the chain initialized with a minsearch algorithm.

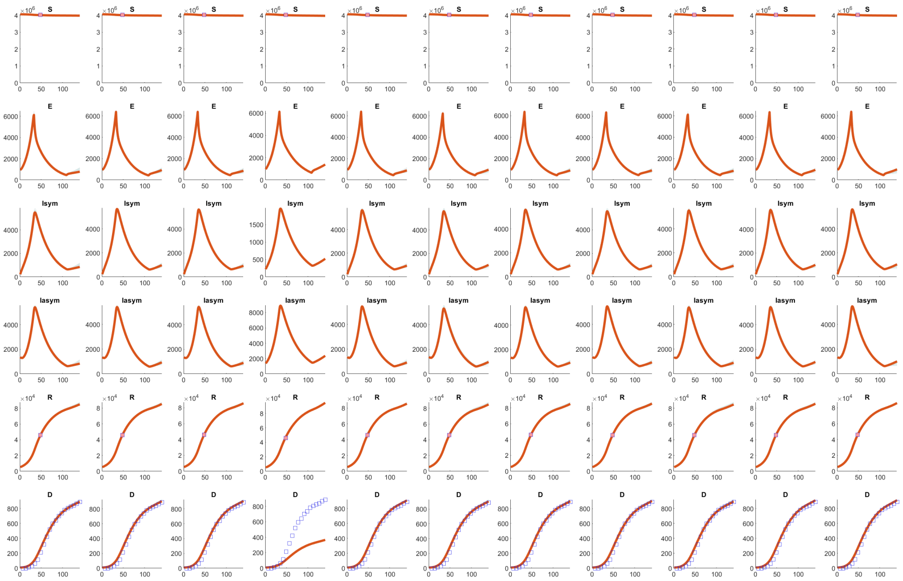


Figure S10: Washington Model Fits. We forward simulate a model using the maximum likelihood estimates of each parameter (red) and 100 random parameter set draws from the posterior distribution (gray). Cumulative deaths are plotted (blue squares), in addition to seroprevalence estimates (orange squares) in each location. Each column corresponds to an MCMC chain, with the first column corresponds to a chain which was initially seeded using a constrained 'minsearch' algorithm. Columns 2-11 correspond to MCMC chains 1-10, which are seeded randomly. Given that chain 3 (column 4) is a poor fit to the data and all other chains seemed to yield reasonable fits, we exclude chain 3 from further analyses.

#### Credible intervals width

The overall width of credible intervals (as shown in the main text) was determined by the value of $c$ (Figure S11). As shown in the trace plots, the value of $c$ was not identifiable from our model simulations, particularly for Washington and New York City, with a wide variety of initial values matching the initial dynamics. In South Florida, the fitted value of $c$ was more constrained, yet model predictions still in large part depended on the ongoing level of social distancing. In part, this issue arose because the relaxation of social distancing in these two locations began after the first wave of the epidemic was largely complete (main text Figure 3), and thus there were few cases shortly after reopening with which to calibrate the dynamics. However, as the outbreak continued, this parameter was crucial for determining the number of deaths by the time shielding was implemented, and hence its ultimate impact. While adding to the length of the time series of deaths used to fit the model might have improved identifiability, ultimately the level of ongoing social distancing is likely to be highly time-varying. The width of the credible interval thus reflects the importance of ongoing social distancing to determine both the trajectory of the United States epidemic and the potential impact of any other control interventions.

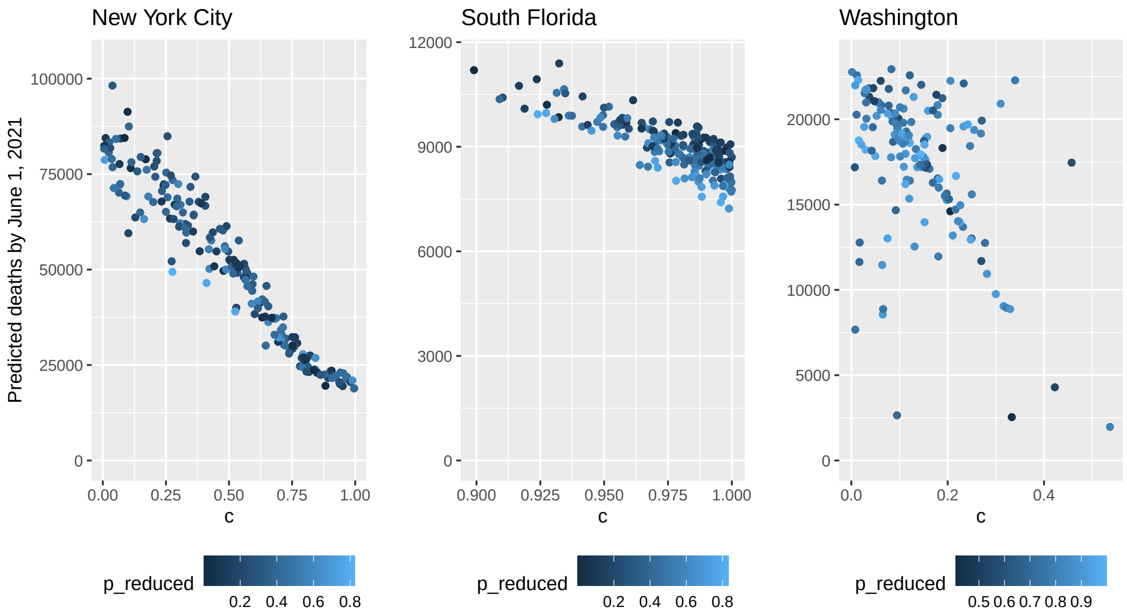


Figure S11: Number of deaths expected after 1 year (y-axis) for each setting (panels) without testing as a function of the degree of relaxation of social distancing (given by $c$, shown on the x-axis) and initial social distancing strength (colors indicate the value of $p_{reduced}$).

### S4. $R_0$ Estimation

The dynamics of the system and $R_0$ are determined by how the outbreak would proceed at time zero in the absence of any interventions, and therefore depends on the fitted values of $p$ and $q$, but not on any of the other fitted parameters. Therefore, we assume no testing at time 0, no social distancing, and no differences in worker contact levels (i.e., all groups mix at the population-average level prior to the outbreak). In this situation, there are only 3 population subgroups at time zero:

$$\frac{dE_{ch}}{dt} = \lambda_{ch} S_{ch} - \gamma_e E_{ch}$$
$$\frac{dI_{s,ch}}{dt} = \gamma_e E_{ch} p - \gamma_s I_{s,ch}$$
$$\frac{dI_{a,ch}}{dt} = \gamma_e E_{ch}(1-p) - \gamma_a I_{a,ch}$$
$$\frac{dH_{s,ch}}{dt} = \gamma_s I_{s,ch}(Hosp_{ch} - CritDie_{ch}) - \gamma_{hs} Hosp_{s,ch}$$
$$\frac{dH_{c,ch}}{dt} = \gamma_s I_{s,ch} CritDie_{ch} - \gamma_{hc} Hosp_{c,ch}$$
$$\frac{dE_{ad}}{dt} = \lambda_{ad} S_{ad} - \gamma_e E_{ad}$$
$$\frac{dI_{s,ad}}{dt} = \gamma_e E_{ad} p - \gamma_s I_{s,ad}$$
$$\frac{dI_{a,ad}}{dt} = \gamma_e E_{ad}(1-p) - \gamma_a I_{a,ad}$$
$$\frac{dH_{s,ad}}{dt} = \gamma_s I_{s,ad}(Hosp_{ad} - CritDie_{ad}) - \gamma_{hs} Hosp_{s,ad}$$
$$\frac{dH_{c,ad}}{dt} = \gamma_s I_{s,ad} CritDie_{ad} - \gamma_{hc} Hosp_{c,ad}$$
$$\frac{dE_{el}}{dt} = \lambda_{el} S_{el} - \gamma_e E_{el}$$
$$\frac{dI_{s,el}}{dt} = \gamma_e E_{el} p - \gamma_s I_{s,el}$$
$$\frac{dI_{a,el}}{dt} = \gamma_e E_{el}(1-p) - \gamma_a I_{a,el}$$
$$\frac{dH_{s,el}}{dt} = \gamma_s I_{s,el}(Hosp_{el} - CritDie_{el}) - \gamma_{hs} Hosp_{s,el}$$
$$\frac{dH_{c,el}}{dt} = \gamma_s I_{s,el} CritDie_{el} - \gamma_{hc} Hosp_{c,el}$$

The $\lambda_i$ for each group is defined as follows.

$$\lambda_{ch} = \frac{qx_{ch,ch}(asyI_{a,ch} + I_{s,ch})}{n_{ch}} + \frac{qx_{ch,ad}(asyI_{a,ad} + I_{s,ad})}{n_{ad}} + \frac{qx_{ch,el}(asyI_{a,el} + I_{s,el})}{n_{el}}$$

$$\lambda_{ad} = \frac{qx_{ad,ch}(asyI_{a,ch} + I_{s,ch})}{n_{ch}} + \frac{qx_{ad,ad}(asyI_{a,ad} + I_{s,ad})}{n_{ad}} + \frac{qx_{ad,el}(asyI_{a,el} + I_{s,el})}{n_{el}}$$

$$\lambda_{el} = \frac{qx_{el,ch}(asyI_{a,ch} + I_{s,ch})}{n_{ch}} + \frac{qx_{el,ad}(asyI_{a,ad} + I_{s,ad})}{n_{ad}} + \frac{qx_{el,el}(asyI_{a,el} + I_{s,el})}{n_{el}}$$

The matrices $\mathcal{F}$ and $\mathcal{V}$ corresponding to these equations are:

$$\mathcal{F} = \begin{bmatrix} 0 & qx_{ch,ch} & qx_{ch,ch}{}^{asy} & 0 & 0 & 0 & qc_{ch,ad} & qx_{ch,ad}{}^{asy} & 0 & 0 & 0 & qx_{ch,el} & qx_{ch,el}{}^{asy} & 0 & 0 \\ 0 & 0 & 0 & 0 & 0 & 0 & 0 & 0 & 0 & 0 & 0 & 0 & 0 & 0 & 0 \\ 0 & 0 & 0 & 0 & 0 & 0 & 0 & 0 & 0 & 0 & 0 & 0 & 0 & 0 & 0 \\ 0 & 0 & 0 & 0 & 0 & 0 & 0 & 0 & 0 & 0 & 0 & 0 & 0 & 0 & 0 \\ 0 & 0 & 0 & 0 & 0 & 0 & 0 & 0 & 0 & 0 & 0 & 0 & 0 & 0 & 0 \\ 0 & qx_{ad,ch} & qx_{ad,ch}{}^{asy} & 0 & 0 & 0 & qx_{ad,ad} & qx_{ad,ad}{}^{asy} & 0 & 0 & 0 & qx_{ad,el} & qx_{ad,el}{}^{asy} & 0 & 0 \\ 0 & 0 & 0 & 0 & 0 & 0 & 0 & 0 & 0 & 0 & 0 & 0 & 0 & 0 & 0 \\ 0 & 0 & 0 & 0 & 0 & 0 & 0 & 0 & 0 & 0 & 0 & 0 & 0 & 0 & 0 \\ 0 & 0 & 0 & 0 & 0 & 0 & 0 & 0 & 0 & 0 & 0 & 0 & 0 & 0 & 0 \\ 0 & 0 & 0 & 0 & 0 & 0 & 0 & 0 & 0 & 0 & 0 & 0 & 0 & 0 & 0 \\ 0 & qx_{el,ch} & qx_{el,ch}{}^{asy} & 0 & 0 & 0 & qx_{el,ad} & qx_{el,ad}{}^{asy} & 0 & 0 & 0 & qx_{el,el} & qx_{el,el}{}^{asy} & 0 & 0 \\ 0 & 0 & 0 & 0 & 0 & 0 & 0 & 0 & 0 & 0 & 0 & 0 & 0 & 0 & 0 \\ 0 & 0 & 0 & 0 & 0 & 0 & 0 & 0 & 0 & 0 & 0 & 0 & 0 & 0 & 0 \\ 0 & 0 & 0 & 0 & 0 & 0 & 0 & 0 & 0 & 0 & 0 & 0 & 0 & 0 & 0 \\ 0 & 0 & 0 & 0 & 0 & 0 & 0 & 0 & 0 & 0 & 0 & 0 & 0 & 0 & 0 \end{bmatrix}$$

$$\mathcal{V} = \begin{bmatrix} \gamma_e & 0 & 0 & 0 & 0 & 0 & 0 & 0 & 0 & 0 & 0 & 0 & 0 & 0 & 0 \\ -\gamma_e p & \gamma_s & 0 & 0 & 0 & 0 & 0 & 0 & 0 & 0 & 0 & 0 & 0 & 0 & 0 \\ -\gamma_e(1-p) & 0 & \gamma_a & 0 & 0 & 0 & 0 & 0 & 0 & 0 & 0 & 0 & 0 & 0 & 0 \\ 0 & -\gamma_s(Hosp_{ch} - Crit_{ch}) & 0 & \gamma_{hs} & 0 & 0 & 0 & 0 & 0 & 0 & 0 & 0 & 0 & 0 & 0 \\ 0 & -\gamma_s Crit_{ch} & 0 & 0 & \gamma_{hc} & 0 & 0 & 0 & 0 & 0 & 0 & 0 & 0 & 0 & 0 \\ 0 & 0 & 0 & 0 & 0 & \gamma_e & 0 & 0 & 0 & 0 & 0 & 0 & 0 & 0 & 0 \\ 0 & 0 & 0 & 0 & 0 & -\gamma_e p & \gamma_s & 0 & 0 & 0 & 0 & 0 & 0 & 0 & 0 \\ 0 & 0 & 0 & 0 & 0 & -\gamma_e(1-p) & 0 & \gamma_a & 0 & 0 & 0 & 0 & 0 & 0 & 0 \\ 0 & 0 & 0 & 0 & 0 & 0 & -\gamma_s(Hosp_{ad} - Crit_{ad}) & 0 & \gamma_{hs} & 0 & 0 & 0 & 0 & 0 & 0 \\ 0 & 0 & 0 & 0 & 0 & 0 & -\gamma_s Crit_{ad} & 0 & 0 & \gamma_{hc} & 0 & 0 & 0 & 0 & 0 \\ 0 & 0 & 0 & 0 & 0 & 0 & 0 & 0 & 0 & 0 & \gamma_e & 0 & 0 & 0 & 0 \\ 0 & 0 & 0 & 0 & 0 & 0 & 0 & 0 & 0 & 0 & -\gamma_e p & \gamma_s & 0 & 0 & 0 \\ 0 & 0 & 0 & 0 & 0 & 0 & 0 & 0 & 0 & 0 & -\gamma_e(1-p) & 0 & \gamma_a & 0 & 0 \\ 0 & 0 & 0 & 0 & 0 & 0 & 0 & 0 & 0 & 0 & 0 & 0 & -\gamma_s(Hosp_{el} - Crit_{el} & \gamma_{hs} & 0 \\ 0 & 0 & 0 & 0 & 0 & 0 & 0 & 0 & 0 & 0 & 0 & 0 & -\gamma_s CritDie_{el} & 0 & \gamma_{hc} \end{bmatrix}$$

To derive an expression for $R_0$ we inverted the $\mathcal{V}$ matrix and multiplied by $\mathcal{F}$. The dominant eigenvalues of this matrix can be computed, but are very complex and are therefore not shown here. It is notable that because hospitalized cases do not contribute to the force of infection, the value of $R_0$ does not depend on $\gamma_{hs}$ or $\gamma_{hc}$. We calculated the value of $R_0$ for each of the three locations based on the fitted values of $p$ and $q$ and the other relevant parameters.

#### S5. Contact matrices

##### Baseline contacts

Baseline contact matrices for 'work' contacts, 'school' contacts, 'home' contacts, and 'other' contacts were taken from [11]. To expand these baseline matrices to the 5 population groups in our model (separating the adult population into $fc$, $rc$, and $h$ classes), we multiplied all contacts with adults $x_{i,ad}$ by the proportion of the adult population falling into each class. We define the fraction of the population falling in each working group as follows:

$$f.home = \frac{n_{home}}{n_{home} + n_{reduced} + n_{full}}$$
$$f.reduced = \frac{n_{reduced}}{n_{home} + n_{reduced} + n_{full}}$$
$$f.full = \frac{n_{full}}{n_{home} + n_{reduced} + n_{full}}$$

For baseline contact matrix values based on [11], we have:

$$x_{i,j} = \begin{bmatrix} x_{ch,ch} & x_{ch,ad} & x_{ch,el} \\ x_{ad,ch} & x_{ad,ad} & x_{ad,el} \\ x_{el,ch} & x_{el,ad} & x_{el,el} \end{bmatrix}$$

For simplicity, we assume that baseline interactions between worker subgroups are only assortative with respect to age (and not with respect to occupation type). To expand this matrix to a 5x5 matrix we use the following notation, where rows 2, 3, and 4 correspond to the work from home, reduced contact, and full contact occupation groups, respectively:

$$x_{i,j} = \begin{bmatrix} x_{ch,ch} & x_{ch,ad}f.home & x_{ch,ad}f.reduced & x_{ch,ad}f.full & x_{ch,el} \\ x_{ad,ch} & x_{ad,ad}f.home & x_{ad,ad}f.reduced & x_{ad,ad}f.full & x_{ch,el} \\ x_{ad,ch} & x_{ad,ad}f.home & x_{ad,ad}f.reduced & x_{ad,ad}f.full & x_{ch,el} \\ x_{ad,ch} & x_{ad,ad}f.home & x_{ad,ad}f.reduced & x_{ad,ad}f.full & x_{ch,el} \\ x_{el,ch} & x_{el,ad}f.home & x_{el,ad}f.reduced & x_{el,ad}f.full & x_{ch,el} \end{bmatrix}$$

Based on these proportions, we define $x_{i,h}$, $x_{i,rc}$, and $x_{i,fc}$ as follows:

$$x_{i,h} = x_{i,ad} f.home$$
$$x_{i,rc} = x_{i,ad} f.reduced$$
$$x_{i,fc} = x_{i,ad} f.full$$

#### Contacts under social distancing

After social distancing has begun, we assume that:

- Home contacts remain the same.
- Schools and daycares close.
- Only working age adults continue to work. Baseline workplace contacts for children and young adults under 20 years of age are nearly zero (average 0.84 contacts/day) and the average workplace contacts for the elderly is 0, so this does not appreciably impact our results.
- All working adults who are able work from home.
- Adults continuing to work outside the home reduce their workplace contacts by constant $p_{reduced}$.
- Other contacts are reduced by scalar constant $sd_{other}$.

The revised contact matrix for work contacts then becomes:

$$CM_{work} = \begin{bmatrix} 0 & 0 & 0 & 0 & 0 \\ 0 & 0 & 0 & 0 & 0 \\ x_{ad,ch}\mathrm{p}_{reduced} & x_{h,h}\mathrm{p}_{reduced} & x_{h,rc}\mathrm{p}_{reduced} & x_{h,fc}\mathrm{p}_{reduced} & x_{ch,el}\mathrm{p}_{reduced} \\ x_{fc,ch} & x_{fc,h} & x_{fc,rc} & x_{fc,fc} & x_{ch,el} \\ 0 & 0 & 0 & 0 & 0 \end{bmatrix}$$

The revised contact matrix for other contacts becomes:

$$CM_{other} = sd.other \times \begin{bmatrix} x_{ch,ch} & x_{ch,h} & x_{ch,rc} & x_{ch,fc} & x_{ch,el} \\ x_{h,ch} & x_{h,h} & x_{h,rc} & x_{h,fc} & x_{ch,el} \\ x_{rc,ch} & x_{rc,h} & x_{rc,rc} & x_{rc,f} & x_{ch,el} \\ x_{fc,ch} & x_{fc,h} & x_{fc,rc} & x_{fc,fc} & x_{ch,el} \\ x_{el,ch} & x_{el,h} & x_{el,rc} & x_{el,fc} & x_{ch,el} \end{bmatrix}$$

#### Contacts during initial relaxing of social distancing

When stay at home orders are initially lifted, we assume that:

- Home contacts remain the same.
- Adults who were working from home continue to work from home.

- Workers in reduced contact occupations increase their workplace contacts based on the intensity of social distancing maintained.
- Schools remain closed until September 1, 2020 in South Florida and until October 1, 2020 in New York City and Washington, after which time they reopen at 50% capacity.
- Other contacts continue to be reduced based on the intensity of social distancing maintained.

#### Contacts after testing begins

When testing begins, we assume that:

- Test-positive individuals move to the test positive group.
- Home contacts remain the same, but their distribution by test status is driven by the proportion of test-positives in the general population.
- Adults who were working from home may return to work if they test positive. Upon returning to work, their workplace contacts are assortative with respect to test status (but not with respect to occupation type).
- Workers in reduced contact occupations increase their workplace contacts based on the intensity of social distancing maintained. Work contacts are preferentially with test-positive individuals, as determined by $\alpha$, or shielding strength.
- Other contacts are increased for test positive individuals to their pre-pandemic levels. Other contacts continue to be reduced for test negative/untested individuals based on the intensity of social distancing maintained. Other contacts are preferentially with test-positive individuals, as determined by $\alpha$, or shielding strength.

After testing has begun, all contact matrices are dependent on the proportion of the population that has tested positive and been released from social distancing at time t. We define this proportion as $r_i(t)$, where $(1 - r_i(t))$ is the fraction of the population who has not yet tested positive.

We assume that social distancing parameters are relaxed from their initial values as follows:

$$p_{reduced} = 1 - (sd_{other} \times c)$$
$$p_{reduced} = 1 - (p_{reduced} \times c)$$

For contact matrices of work and 'other' contacts, we implement shielding factor $\alpha$, which increases the probability of contacting a test-positive individual according to their prevalence in the population (achieved by multiplying expected contact rates due to prevalence by scaling factor $\alpha + 1$). To account for the fact that, when prevalence is high, $(\alpha + 1)r_i(t)$ may exceed 1, we introduce a variable $s_i(t)$:

$$s_i(t) = \begin{cases} (\alpha + 1)r_i(t) & (\alpha + 1)r_i(t) \leq 1 \\ 1 & (\alpha + 1)r_i(t) \geq 1 \end{cases}$$

This shielding structure is similar to ‘fixed shielding’, previously described by Weitz et al [18] in that it preserves the baseline number of contacts and increases contacts for test positive individuals by $1 + \alpha$, as shown below:

$$
\begin{aligned}
x_0 &= x_0 r_i(t) + x_0(1 - r_i(t)) \\
x_0 &= r_i(t)(\alpha + 1)x_0 + (x_0 - (\alpha + 1)r_i(t)x_0)
\end{aligned}
$$

The structure of all three matrices (home, work, and other) is given by CM:

$$CM_{home} = \begin{bmatrix} r_{ch}(t)x_{ch,ch} & (1-r_{ch}(t))x_{ch,ch} & r_h(t)x_{ch,h} & (1-r_h(t))x_{ch,h} & r_{rc}(t)x_{ch,rc} & (1-r_{rc}(t))x_{ch,rc} & r_{fc}(t)x_{ch,fc} & (1-r_{fc}(t))x_{ch,fc} & r_{el}(t)x_{ch,el} & (1-r_{el}(t))x_{ch,el} \\ r_{ch}(t)x_{ch,ch} & (1-r_{ch}(t))x_{ch,ch} & r_h(t)x_{ch,h} & (1-r_h(t))x_{ch,h} & r_{rc}(t)x_{ch,rc} & (1-r_{rc}(t))x_{ch,rc} & r_{fc}(t)x_{ch,fc} & (1-r_{fc}(t))x_{ch,fc} & r_{el}(t)x_{ch,el} & (1-r_{el}(t))x_{ch,el} \\ r_{ch}(t)x_{h,ch} & (1-r_{ch}(t))x_{h,ch} & r_h(t)x_{h,h} & (1-r_h(t))x_{h,h} & r_{rc}(t)x_{h,rc} & (1-r_{rc}(t))x_{h,rc} & r_{fc}(t)x_{h,fc} & (1-r_{fc}(t))x_{h,fc} & r_{el}(t)x_{h,el} & (1-r_{el}(t))x_{h,el} \\ r_{ch}(t)x_{h,ch} & (1-r_{ch}(t))x_{h,ch} & r_h(t)x_{h,h} & (1-r_h(t))x_{h,h} & r_{rc}(t)x_{h,rc} & (1-r_{rc}(t))x_{h,rc} & r_{fc}(t)x_{h,fc} & (1-r_{fc}(t))x_{h,fc} & r_{el}(t)x_{h,el} & (1-r_{el}(t))x_{h,el} \\ r_{ch}(t)x_{rc,ch} & (1-r_{ch}(t))x_{rc,ch} & r_h(t)x_{rc,h} & (1-r_h(t))x_{rc,h} & r_{rc}(t)x_{rc,rc} & (1-r_{rc}(t))x_{rc,rc} & r_{fc}(t)x_{rc,fc} & (1-r_{fc}(t))x_{rc,fc} & r_{el}(t)x_{rc,el} & (1-r_{el}(t))x_{rc,el} \\ r_{ch}(t)x_{rc,ch} & (1-r_{ch}(t))x_{rc,ch} & r_h(t)x_{rc,h} & (1-r_h(t))x_{rc,h} & r_{rc}(t)x_{rc,rc} & (1-r_{rc}(t))x_{rc,rc} & r_{fc}(t)x_{rc,fc} & (1-r_{fc}(t))x_{rc,fc} & r_{el}(t)x_{rc,el} & (1-r_{el}(t))x_{rc,el} \end{bmatrix}$$

$$CM_{other} = \begin{bmatrix} 1 \\ sd.other2 \\ 1 \\ sd.other2 \\ 1 \\ sd.other2 \\ 1 \\ sd.other2 \\ 1 \\ sd.other2 \end{bmatrix} \times \begin{bmatrix} s_{ch}(t)x_{ch,ch} & (1-s_{ch}(t))x_{ch,ch} & s_h(t)x_{ch,h} & (1-s_h(t))x_{ch,h} & s_{rc}(t)x_{ch,rc} & (1-s_{rc}(t))x_{ch,rc} & s_{fc}(t)x_{ch,fc} & (1-s_{fc}(t))x_{ch,fc} & s_{el}(t)x_{ch,el} & (1-s_{el}(t))x_{ch,el} \\ s_{ch}(t)x_{ch,ch} & (1-s_{ch}(t))x_{ch,ch} & s_h(t)x_{ch,h} & (1-s_h(t))x_{ch,h} & s_{rc}(t)x_{ch,rc} & (1-s_{rc}(t))x_{ch,rc} & s_{fc}(t)x_{ch,fc} & (1-s_{fc}(t))x_{ch,fc} & s_{el}(t)x_{ch,el} & (1-s_{el}(t))x_{ch,el} \\ s_{ch}(t)x_{h,ch} & (1-s_{ch}(t))x_{h,ch} & s_h(t)x_{h,h} & (1-s_h(t))x_{h,h} & s_{rc}(t)x_{h,rc} & (1-s_{rc}(t))x_{h,rc} & s_{fc}(t)x_{h,fc} & (1-s_{fc}(t))x_{h,fc} & s_{el}(t)x_{h,el} & (1-s_{el}(t))x_{h,el} \\ s_{ch}(t)x_{h,ch} & (1-s_{ch}(t))x_{h,ch} & s_h(t)x_{h,h} & (1-s_h(t))x_{h,h} & s_{rc}(t)x_{h,rc} & (1-s_{rc}(t))x_{h,rc} & s_{fc}(t)x_{h,fc} & (1-s_{fc}(t))x_{h,fc} & s_{el}(t)x_{h,el} & (1-s_{el}(t))x_{h,el} \\ s_{ch}(t)x_{rc,ch} & (1-s_{ch}(t))x_{rc,ch} & s_h(t)x_{rc,h} & (1-s_h(t))x_{rc,h} & s_{rc}(t)x_{rc,rc} & (1-s_{rc}(t))x_{rc,rc} & s_{fc}(t)x_{rc,fc} & (1-s_{fc}(t))x_{rc,fc} & s_{el}(t)x_{rc,el} & (1-s_{el}(t))x_{rc,el} \\ s_{ch}(t)x_{rc,ch} & (1-s_{ch}(t))x_{rc,ch} & s_h(t)x_{rc,h} & (1-s_h(t))x_{rc,h} & s_{rc}(t)x_{rc,rc} & (1-s_{rc}(t))x_{rc,rc} & s_{fc}(t)x_{rc,fc} & (1-s_{fc}(t))x_{rc,fc} & s_{el}(t)x_{rc,el} & (1-s_{el}(t))x_{rc,el} \\ s_{ch}(t)x_{fc,ch} & (1-s_{ch}(t))x_{fc,ch} & s_h(t)x_{fc,h} & (1-s_h(t))x_{fc,h} & s_{rc}(t)x_{fc,rc} & (1-s_{rc}(t))x_{fc,rc} & s_{fc}(t)x_{fc,fc} & (1-s_{fc}(t))x_{fc,fc} & s_{el}(t)x_{fc,el} & (1-s_{el}(t))x_{fc,el} \\ s_{ch}(t)x_{fc,ch} & (1-s_{ch}(t))x_{fc,ch} & s_h(t)x_{fc,h} & (1-s_h(t))x_{fc,h} & s_{rc}(t)x_{fc,rc} & (1-s_{rc}(t))x_{fc,rc} & s_{fc}(t)x_{fc,fc} & (1-s_{fc}(t))x_{fc,fc} & s_{el}(t)x_{fc,el} & (1-s_{el}(t))x_{fc,el} \\ s_{ch}(t)x_{el,ch} & (1-s_{ch}(t))x_{el,ch} & s_h(t)x_{el,h} & (1-s_h(t))x_{el,h} & s_{rc}(t)x_{el,rc} & (1-s_{rc}(t))x_{el,rc} & s_{fc}(t)x_{el,fc} & (1-s_{fc}(t))x_{el,fc} & s_{el}(t)x_{el,el} & (1-s_{el}(t))x_{el,el} \\ s_{ch}(t)x_{el,ch} & (1-s_{ch}(t))x_{el,ch} & s_h(t)x_{el,h} & (1-s_h(t))x_{el,h} & s_{rc}(t)x_{el,rc} & (1-s_{rc}(t))x_{el,rc} & s_{fc}(t)x_{el,fc} & (1-s_{fc}(t))x_{el,fc} & s_{el}(t)x_{el,el} & (1-s_{el}(t))x_{el,el} \end{bmatrix}$$

$$CM_{work} = \begin{bmatrix} 0 \\ 0 \\ 1 \\ 0 \\ preduced2 \\ preduced2 \\ 1 \\ 1 \\ 0 \\ 0 \end{bmatrix} \times \begin{bmatrix} 0 & 0 & 0 & 0 & 0 & 0 & 0 & 0 & 0 & 0 \\ 0 & 0 & 0 & 0 & 0 & 0 & 0 & 0 & 0 & 0 \\ x_{h,ch} & 0 & x_{h,h} & 0 & x_{h,rc} & 0 & x_{h,fc} & 0 & x_{h,el} & 0 \\ 0 & 0 & 0 & 0 & 0 & 0 & 0 & 0 & 0 & 0 \\ s_{ch}(t)x_{rc,ch} & (1-s_{ch}(t))x_{rc,ch} & s_h(t)x_{rc,h} & (1-s_h(t))x_{rc,h} & s_{rc}(t)x_{rc,rc} & (1-s_{rc}(t))x_{rc,rc} & s_{fc}(t)x_{rc,fc} & (1-s_{fc}(t))x_{rc,fc} & s_{el}(t)x_{rc,el} & (1-s_{el}(t))x_{rc,el} \\ s_{ch}(t)x_{rc,ch} & (1-s_{ch}(t))x_{rc,ch} & s_h(t)x_{rc,h} & (1-s_h(t))x_{rc,h} & s_{rc}(t)x_{rc,rc} & (1-s_{rc}(t))x_{rc,rc} & s_{fc}(t)x_{rc,fc} & (1-s_{fc}(t))x_{rc,fc} & s_{el}(t)x_{rc,el} & (1-s_{el}(t))x_{rc,el} \\ 0 & 0 & 0 & 0 & 0 & 0 & 0 & 0 & 0 & 0 \\ 0 & 0 & 0 & 0 & 0 & 0 & 0 & 0 & 0 & 0 \end{bmatrix}$$

#### S6. Overall contact reductions by location

| Location | Reduction in work contacts for adults | Reduction in other contacts for all groups | Reduction in total contacts Children | Adults | Elderly |
|---|---|---|---|---|---|
| New York City | 52.3% | 43.9% | 33.8% | 37.6% | 25.5% |
| South Florida | 70.2% | 78.4% | 45.8% | 56.1% | 44.8% |
| Washington | 32.1% | 10.5% | 21.9% | 18.6% | 6.9% |

Table S3: Steady state reduction in contacts by location after stay-at-home orders are lifted and schools reopen
